# Real-world comparative effectiveness of a third dose of mRNA-1273 versus BNT162b2 among adults aged ≥65 years in the United States

**DOI:** 10.1101/2023.11.03.23298054

**Authors:** Brenna Kirk, Chris Bush, Astra Toyip, Katherine E. Mues, Ekkehard Beck, Linwei Li, Samantha St. Laurent, Mihaela Georgieva, Morgan A. Marks, Tianyu Sun, Daina B. Esposito, David Martin, Nicolas Van de Velde

**Affiliations:** Aetion, Inc., 5 Pennsylvania Plaza, New York, NY 10001, USA; Moderna, Inc., 200 Technology Square, Cambridge, MA 02139, USA

**Keywords:** Booster, COVID-19, mRNA-1273, Vaccine Effectiveness

## Abstract

**Introduction:** To compare the real-world effectiveness of a third dose of mRNA-1273 versus a third dose of BNT162b2 against breakthrough COVID-19 hospitalizations among adults age ≥65 years who completed a primary series of an mRNA-based COVID-19 vaccine (regardless of which primary series was received).

**Materials and methods:** This observational comparative vaccine effectiveness (VE) study was conducted using administrative claims data from the US HealthVerity database (September 22, 2021, to August 31, 2022). A third dose of mRNA-1273 versus BNT162b2 was assessed for preventing COVID-19 hospitalizations and medically attended COVID-19 among adults ≥65 years. Inverse probability of treatment weighting was applied to balance baseline characteristics between vaccine groups. Incidence rates from patient-level data and hazard ratios (HRs) with 95% confidence intervals (CIs) using weighted Cox proportional hazards models were calculated to estimate relative VE for each outcome.

**Results:** Overall, 94,587 and 92,377 individuals received a third dose of mRNA-1273 and BNT162b2, respectively. Among the weighted population, the median age was 69 years (interquartile range, 66-74), 53% were female, and 46% were commercially insured. COVID-19 hospitalization rates per 1000 person-years (PYs) were 5.61 (95% CI, 5.13-6.09) for mRNA-1273 and 7.06 (95% CI, 6.54-7.57) for BNT162b2 (HR, 0.82; 0.69-0.98). Medically attended COVID-19 rates per 1000 PYs (95% CI) were 95.05 (95% CI, 93.03-97.06) for mRNA-1273 and 106.55 (95% CI, 104.53-108.57) for BNT162b2 (HR, 0.93; 0.89-0.98).

**Conclusions:** Results from this observational comparative VE database study provide evidence that among older adults, a third dose of mRNA-1273 was more effective in preventing breakthrough COVID-19 hospitalization and medically attended COVID-19 infection compared with a third dose of BNT162b2.

## 1. Introduction

As of July 19, 2023, there have been more than 103.4 million confirmed cases of COVID-19 in the United States, with 1,127,152 deaths [1]. The development and widespread distribution of efficacious vaccines against COVID-19 is estimated to have saved nearly 20 million lives between December 2020 and December 2021 [2]. Therefore, continuing to evaluate the performance of these vaccines is of pertinent concern to the evolving public health landscape, particularly as the SARS-CoV2 virus continues to evolve.

The Pfizer-BioNTech (BNT162b2) COVID-19 vaccine, also known as COMIRNATY, was the first COVID-19 vaccine to be fully approved by the US Food and Drug Administration (FDA) on August 23, 2021, as a 2-dose primary series consisting of 2 injections administered 21 days apart [3]. More recently, vaccination guidelines have been updated to include additional booster doses to combat waning vaccine efficacy over time and to address the emergence of new COVID-19 strains [4]. The initial phase 2/3 clinical trials in participants aged ≥16 years who received booster doses demonstrated a 95% effectiveness in preventing COVID-19 infection [5]. On September 22, 2021, the FDA provided emergency use authorization (EUA) for the administration of BNT162b2 as a booster dose ≥6 months after primary series completion. On November 19, 2021, the FDA approved the use of a single booster dose of BNT162b2 for administration to all individuals ≥18 years after completion of primary series with any FDA-authorized or approved COVID-19 vaccine [6].

The Moderna COVID-19 vaccine (mRNA-1273), or Spikevax, is authorized as a 2-dose primary series to be given as 2 injections 28 days apart for immunocompetent individuals aged ≥12 years; similar to BNT-162b2, an additional (third) dose of mRNA-1273 is recommended for immunocompromised (IC) patients [7]. The phase 3 clinical trials demonstrated a 94.1% vaccine efficacy for COVID-19 symptomatic illness [8]. A booster dose following the primary series of mRNA-1273 was approved via an EUA on October 20, 2021 [7]. On August 31, 2022, the FDA authorized both the Pfizer and Moderna omicron-containing bivalent COVID-19 booster vaccines under an EUA [7].

To date, several studies have indicated a greater vaccine efficacy for mRNA-1273 compared with BNT162b2 in populations who have completed a primary vaccine series [9], as well as in those who have received an additional vaccine dose [10–15]. One study conducted with data from the US Department of Veterans Affairs utilized a test-negative case-control design to demonstrate a primary series vaccine effectiveness (VE) of 96.2% (95% CI, 95.5%-96.9%) for recipients of BNT162b2, and 98.2% (95% CI, 97.5%-98.6%) for mRNA-1273 against COVID-19 infection [9]. A retrospective cohort study emulating a clinical trial and utilizing electronic health records to estimate comparative effectiveness of a third dose found the excess number of COVID-19 infections over 9 weeks for BNT162b2 compared to mRNA-1273 was 63.2 per 10,000 persons (95% CI, 15.2-100.7) [15]. Another study, using the OpenSAFELY-TPP database, assessed the comparative effectiveness of booster doses and third primary series doses of BNT162b2 versus mRNA-1273 in England. When comparing mRNA-1273 to BNT162b2, the authors calculated a hazard ratio (HR) of 0.95 (95% CI, 0.95-0.96) against COVID-19 infection and 0.89 (95% CI, 0.82-0.95) for COVID-19 hospitalization [14]. Outside of the United States, a retrospective cohort study conducted using a Japanese registry found that patients with an mRNA-1273 booster dose had a lower risk of COVID-19 infection than those with a BNT162b2 booster (HR, 0.62; 95% CI, 0.50-0.74) [12].

Adults aged ≥65 years are at an increased risk of severe disease, hospitalization, and death as a result of COVID-19 [16]. Additionally, a significant proportion of older adults suffer from conditions such as hypertension, cardiovascular disease, diabetes, chronic respiratory disease, and chronic kidney disease, which have all been associated with increased COVID-19–related morbidity and mortality [17]. While the vaccines have been effective in preventing severe disease and death in this population, a study by Newman et al showed diminished activity of neutralizing antibodies in patients aged 80 to 89 years vaccinated with BNT162b2 compared with patients aged 70 to 79 years, suggesting that vaccine efficacy may decline with age [18].

There remains interest in the use and effectiveness of mRNA vaccines among older adults. A study by Tenforde et al compared adults aged ≥65 years who were partially or fully vaccinated with either BNT162b2 or mRNA-1273 to those who were unvaccinated, estimating VE against COVID-19 of 94% (95% CI, 49%-99%) among fully vaccinated individuals and 64% (95% CI, 28%-82%) among partially vaccinated individuals [19]. However, to date, there is limited research on the relative real-world VE of a third dose (booster or third primary series) between the available mRNA-based COVID-19 vaccines in individuals aged ≥65 years. The updated booster dose guidelines, along with the prevalence of new strains and waning vaccine efficacy with age and time, underscore the significance of this research. The present study generated real-world evidence on the comparative VE of 3-dose vaccine series in individuals aged ≥65 years to inform healthcare practitioners (HCPs) and public health decision-making.

## 2. Methods

### 2.1 Objectives

To compare the real-world VE of a third dose of mRNA-1273 versus a third dose of BNT162b2 against breakthrough COVID-19 hospitalization and medically attended COVID-19 among adults aged ≥65 years who completed a primary series of an mRNA-based COVID-19 vaccine (regardless of which primary series was received).

### 2.2 Study design and data

The HealthVerity database has been previously described [20]. Briefly, the database includes demographic variables such as age, sex, and 3-digit zip code level. Hospitalizations are included in the data at a summary level. Open claims are sourced directly through medical clearinghouses and pharmacy benefit managers; there is no associated enrollment file, resulting in incomplete capture of healthcare system interactions. Closed claims represent claims accepted by and paid by health insurance companies and generally lagged 3 to 6 months to allow for full adjudication. Both open and closed claims were drawn from a variety of US sources, including a closed claims database, Private Source 20 (PS20). The sample of patients from PS20 was enriched with open and closed medical and pharmacy claims feeds linked through HealthVerity, including Private Source 17 (PS17), which includes adjudicated pharmacy claims sourced from a pharmacy benefit manager and an associated pharmacy enrollment file. For a proportion of patients in PS17 (approximately 23%), adjudicated medical claims are linked to the pharmacy enrollment file. For patients with linked medical claims in PS17, 100% of adjudicated medical claims (all claims for those patients) will be observable over the time a patient is enrolled in the pharmacy benefit manager plan.

This retrospective comparative VE cohort study utilized medical and pharmacy claims data aggregated by HealthVerity from September 22, 2021 (when booster vaccine doses were first approved) through August 31, 2022. The primary analysis was limited to closed medical claims only from PS20 for inclusion/exclusion criteria as well as covariates, with outcomes assessed in either open or closed claims data. Data were truncated August 31, 2022, to ensure complete capture of all medical and pharmacy records.

The study follows the Guidelines for Good Epidemiologic Practice laid out in the 2005 US FDA Good Pharmacy Practice and the 2008 International Society of Pharmacoepidemiology Good Pharmacy Practice. This study was fully approved by the WCG Institutional Review Board (#20231679). All participant data were de-identified, and all participant-level and provider-level data within the database contained synthetic identifiers to protect the privacy of individuals and data contributors.

### 2.3 Population and follow-up

Individuals aged ≥65 years who received 3 doses of mRNA-1273 or BNT162b2 and were continuously enrolled in a medical and pharmacy plan for 365 days before the index date were included in the analysis (Figure 1). The index date is defined as a patient’s first qualifying vaccine claim for a third dose of either the mRNA-1273 or BNT162b2 vaccine identified between September 22, 2021, through August 31, 2022 (Figure 1). The third dose must have occurred after September 22, 2021, and at least 42 days following completion of a second dose in a 2-dose primary mRNA vaccine series. A second dose of mRNA-1273 was required to occur within 23 to 33 days (28 days ± 5 days) following a first dose of mRNA-1273, and a second dose of BNT162b2 was required to occur 16 to 26 days (21 days ± 5 days) following a first dose of BNT162b2. A second dose in a heterologous primary vaccine series was required to occur 23 to 33 days following a first dose. Vaccinations were captured in the database via manufacturer-specific current procedural terminology (CPT) and national drug code (NDC) (Table S1). See Supplementary File 1 for additional details on the data-driven decision for timing between first and second doses as well as timing for the third dose. The baseline period was defined as the 365 days before the index date. All patients were required to have continuous medical and pharmacy claims enrollment with no allowable gap during the baseline period. Patients were excluded if they had missing age or sex on index, evidence of a prior COVID-19 infection any time before the index date, or evidence of administration of a non–mRNA COVID-19 vaccine any time before the index date. Follow-up began 14 days following the index date, in order to allow patients to reach full levels of immunogenicity following vaccination. Patients who disenrolled, reached the end of their available data, had a COVID-19 diagnosis or COVID-19 vaccine event between day 1 and day 13 following the index date were also excluded. Participants were followed until an outcome of interest, receipt of a fourth COVID-19 vaccine dose (any manufacturer), disenrollment from a medical/pharmacy plan, or August 31, 2022, whichever occurred first.

**Figure 1.**
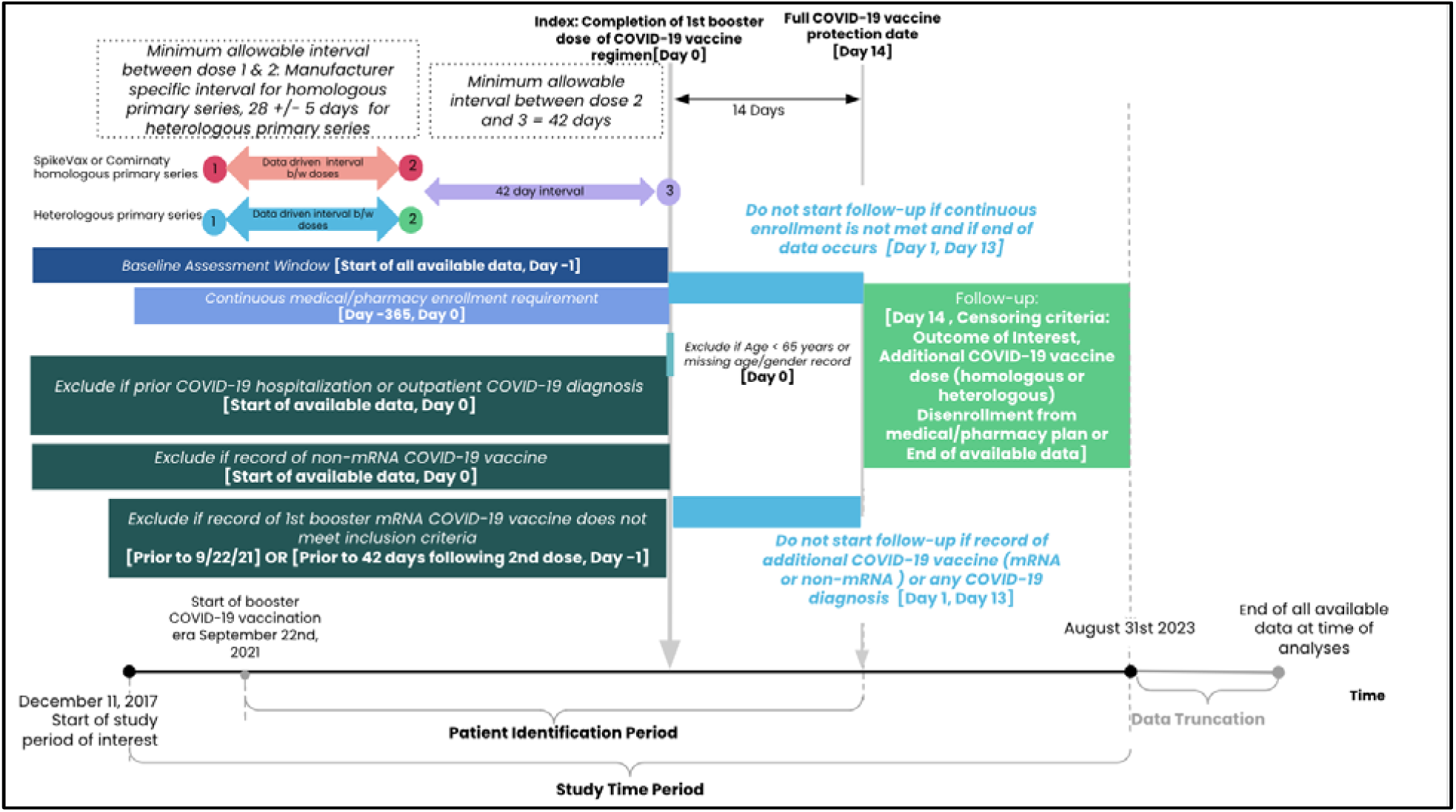
Study design diagram.

### 2.4 Study outcomes

The primary outcome of interest in this study was a COVID-19 hospitalization defined as a hospitalization episode overlapping with an International Statistical Classification of Diseases and Related Health Problems, 10^th^ revision (ICD-10) diagnosis code for COVID-19 (U07.1) in any position, any setting (inpatient, outpatient, emergency department, urgent care, etc.), and an ICD-10 diagnosis code indicating evidence of respiratory distress in any position. Because HealthVerity does not combine individual inpatient claims into a single hospitalization, a hospitalization episode was defined as concurrent/adjacent hospital claims with a 4-day allowable gap. Supplementary File 2 provides details on the rationale for how this hospitalization was defined. A respiratory distress diagnosis was based on conditions used in Kluberg *et al* and Garry *et al* and included unspecified coronavirus infection, pneumonia, pneumonia due to SARS-associated coronavirus, acute respiratory distress syndrome, sepsis, bronchitis, cough, shortness of breath, diagnosis indicating failed intubation, hypoxemia, supplemental O_2_ use, respiratory infection, or ventilation-related diagnosis codes [21, 22]. The secondary outcome of interest in this study was medically attended COVID, which was defined as a medical claim with the ICD-10 diagnostic code for COVID-19 (U07.1) in any setting, including inpatient, outpatient, emergency department, and urgent care.

### 2.5 Statistical analysis

The distribution of baseline variables within each vaccine group was described as number and percentage for categorical variables and as mean (standard deviation), median (interquartile range), and range (minimum, maximum) for continuous variables. Baseline variables hypothesized to be confounders were included in a propensity score model. Age, sex, payer type, state of residence, and calendar time (year and month) were assessed at index. Healthcare resource utilization, Charlson comorbidity score, and frailty score (from a claims-based frailty index developed by Kim et al [23]) were assessed in the 365 days before the index date. Individual comorbid conditions were assessed using all prior claims available. The full list of baseline covariates included in the propensity score model is presented in Table S2.

Inverse probability of treatment weights was calculated as 1/PS for participants in the mRNA-1273 group (exposed) and as 1/(1-PS) for participants in the BNT162b2 group (referent). Weights were truncated at the 99^th^ percentile in order to account for large weights resulting from patients in the exposure group who had a low propensity for being exposed. Exposure and referent groups were considered balanced if the absolute standardized differences (ASDs) for all baseline covariates used to generate the PS were <0.10 [24, 25]. Incidence rates per 1000 PYs with corresponding 95% confidence intervals (CIs) were estimated for each treatment group. Weighted Cox proportional hazard models were used to estimate HRs and corresponding 95% CIs. Kaplan–Meier plots with 95% CIs and Schoenfeld residuals were generated to assess the proportional hazards assumption.

The following subgroup analyses were implemented: primary vaccine series (3-dose homologous series, homologous 2-dose series of mRNA-1273, homologous 2-dose series of BNT162b2, and heterologous primary series), calendar quarter of vaccine receipt, and IC status. IC status was defined in accordance with the definition reported by Mues et al [20] and is further outlined in Supplement Table S3. In order to best capture balance within the primary vaccine series subgroups, the PS was re-calculated within each of these subgroups.

Analyses were conducted using the Aetion Evidence Platform^®^ software for real-world data analysis, which has been scientifically validated for observational cohort studies using large healthcare databases [26]. Transformations of the raw data are preserved for full reproducibility, and audit trails are available, including a quality check of the data ingestion process. Kaplan– Meyer plots were produced using R version 4.3.1.

### 2.6 Sensitivity analysis

Several per-protocol sensitivity analyses were conducted. First, two alternative definitions of COVID-19 hospitalization were implemented: 1) required an inpatient claim with an ICD-10 diagnosis of COVID-19 in any diagnosis position and no evidence of respiratory distress and 2) a hospitalization episode overlapping with a COVID-19 diagnosis and a respiratory distress diagnosis where the codes to identify respiratory distress were more restrictive than the definition applied in the primary analysis (see Supplementary file S3). The code list was modified from Kluberg et al [21]. Second, the impact of booster vaccine–specific exclusion criteria was assessed by not excluding patients based on receiving a third COVID-19 vaccine dose occurring before September 22, 2021 (date of the EUA for the BNT162b2 booster vaccine), or within 41 days following the completion of a primary series. Third, the impact of inclusion of open claims into patient inclusion criteria and covariate identification was assessed.

With covariates and index variables being captured with open or closed claims, patients were required to have 365 days of continuous baseline enrollment in order to ensure observability among the patients captured. A final sensitivity analysis was conducted to assess the impact of modifying the window for COVID-19 exclusion at baseline from all available data to the 180 days before the index date.

## 3. Results

There were 186,964 adults ≥65 years who met the study inclusion criteria between September 22, 2021, and August 31, 2022, of which 94,587 (50.6%) and 92,377 (49.4%) received a third dose of mRNA-1273 and BNT162b2, respectively (Figure 2). There were fewer commercially insured adults among those administered mRNA-1273 compared with BNT162b2 (42.7% vs 48.4%, respectively), while age and sex distributions were comparable between the groups prior to weighting (Table 1). In the pre-weighted cohort, In September 2021, <1% of adults had received a third dose of mRNA-1273 while 8.8% of adults received a third dose of BNT162b2.

**Figure 2.**
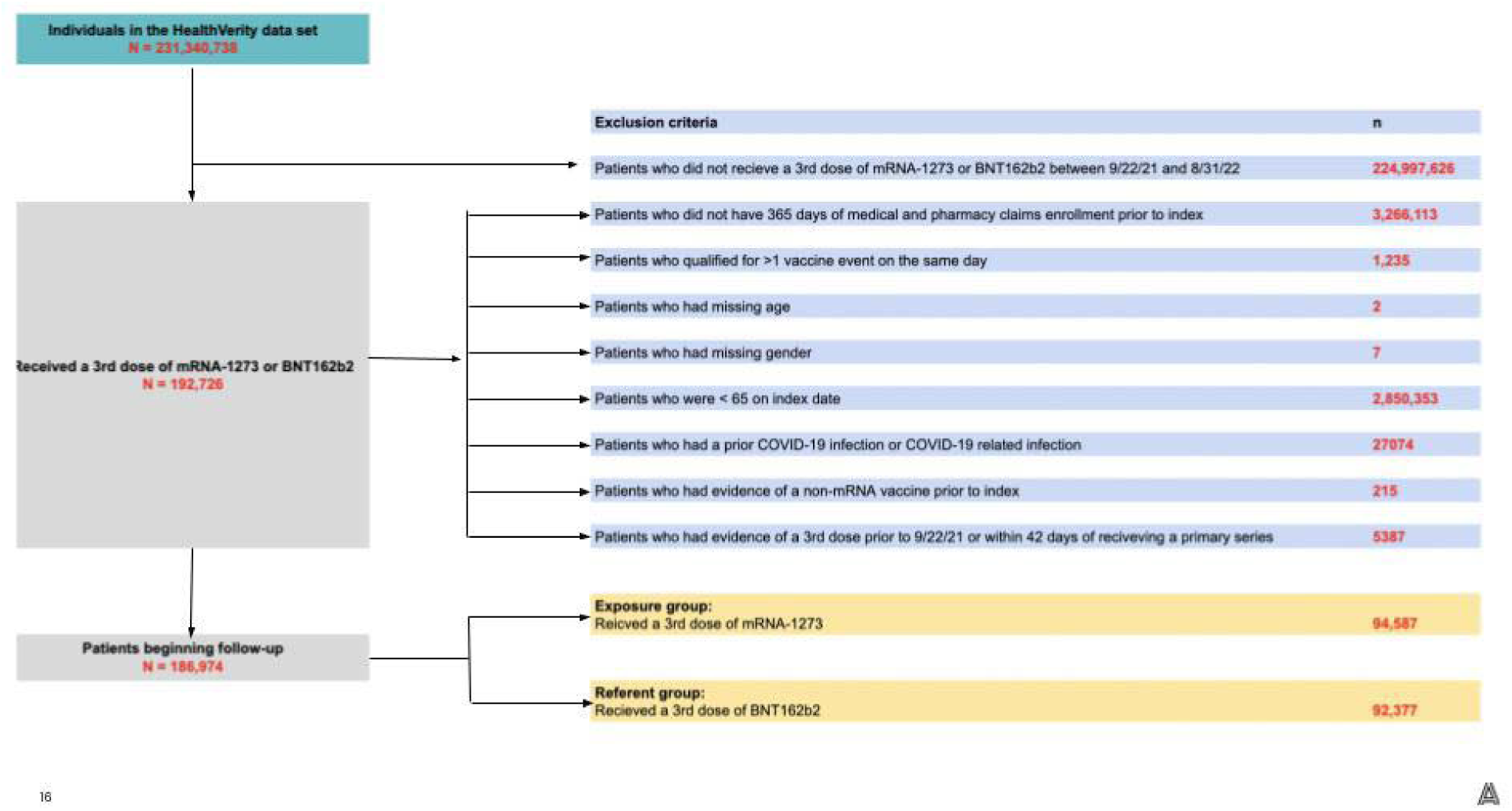
CONSORT diagram.

**Table 1.** Pre- and post-PS weighting baseline characteristics for primary population.

|  | Pre-PS weighting baseline characteristics |  |  | Post-PS weighting (truncated at 99th percentile) |  |  |
| --- | --- | --- | --- | --- | --- | --- |
|  | Third dose of mRNA-1273 | Third dose of BNT162b2 | Absolute standardized differences | Third dose of mRNA-1273 | Third dose of BNT162b2 | Weighted absolute standardized difference |
| **Number of patients** | 94,587 | 92,377 |  | 94,587 | 92,377 |  |
| Sum of weights |  |  |  | 180,606.89 | 186,618.19 |  |
| Effective sample size |  |  |  | 85,575 | 81,996 |  |
| **Age (continuous)** |  |  | 0.004 |  |  | 0.003 |
| ...Mean (SD), years | 71.63 (7.76) | 71.59 (8.03) |  | 71.46 (7.76) | 71.48 (7.83) |  |
| Median [IQR], years | 69.00 [66.00, 74.00] | 69.00 [66.00, 74.00] |  | 69.00 [66.00, 74.00] | 69.00 [66.00, 74.00] |  |
| **Age (categorical)** |  |  |  |  |  |  |
| ...65-74 years | 72,076 (76.2%) | 70,451 (76.3%) | 0.002 | 138,833.9 (76.9%) | 143,387.9 (76.8%) | 0.001 |
| ...85-84 years | 15,486 (16.4%) | 14,405 (15.6%) | 0.021 | 28,417.7 (15.7%) | 29,249.8 (15.7%) | 0.002 |
| ...85+ years | 7,025 (7.4%) | 7,521 (8.1%) | 0.027 | 13,355.3 (7.4%) | 13,980.5 (7.5%) | 0.004 |
| **Sex** |  |  |  |  |  |  |
| ...Female | 50,636 (53.5%) | 49,511 (53.6%) | 0.001 | 96,717.0 (53.6%) | 99,774.9 (53.5%) | 0.002 |
| ...Male | 43,951 (46.5%) | 42,866 (46.4%) | 0.001 | 83,889.9 (46.5%) | 86,843.3 (46.5%) | 0.002 |
| **Primary payer type** |  |  |  |  |  |  |
| ...Missing | 3143 (3.3%) | 4528 (4.9%) | 0.08 | 6879.9 (3.8%) | 7489.5 (4.0%) | 0.011 |
| ...Commercial | 40,377 (42.7%) | 44,708 (48.4%) | 0.115 | 82,593.9 (45.7%) | 86,008.4 (46.1%) | 0.007 |
| ...Medicare | 27,874 | 23,387 | 0.093 | 49,369.4 | 50,584.8 | 0.005 |
|  | (29.5%) | (25.3%) |  | (27.3%) | (27.1%) |  |
| ...Medicaid | 23,193<br>(24.5%) | 19,754<br>(21.4%) | 0.075 | 41,763.6<br>(23.1%) | 42,535.4<br>(22.8%) | 0.008 |
| **State** |  |  |  |  |  |  |
| ...Alaska/Other/<br>PR | 132 (0.1%) | 49 (0.1%) | 0.028 | 177.4 (0.1%) | 164.6 (0.1%) | 0.003 |
| ...Alabama | 94 (0.1%) | 82 (0.1%) | 0.003 | 174.7 (0.1%) | 176.3 (0.1%) | 0.001 |
| ...Arkansas | 2415 (2.6%) | 2422 (2.6%) | 0.004 | 4561.4<br>(2.5%) | 4792.9<br>(2.6%) | 0.003 |
| ...Arizona | 13,262<br>(14.0%) | 6028 (6.5%) | 0.249 | 19,096.6<br>(10.6%) | 18,864.2<br>(10.1%) | 0.015 |
| ...California | 10,438<br>(11.0%) | 6091 (6.6%) | 0.157 | 16,349.6<br>(9.0%) | 16,587.3<br>(8.9%) | 0.006 |
| ...Colorado | 280 (0.3%) | 313 (0.3%) | 0.008 | 567.8 (0.3%) | 589.7 (0.3%) | 0.000 |
| ...Connecticut | 520 (0.5%) | 857 (0.9%) | 0.044 | 1334.9<br>(0.7%) | 1380.5<br>(0.7%) | 0.000 |
| ...District of<br>Columbia | 179 (0.2%) | 134 (0.1%) | 0.011 | 313.1 (0.2%) | 322.7 (0.2%) | 0.000 |
| ...Delaware | 302 (0.3%) | 467 (0.5%) | 0.029 | 711.9 (0.4%) | 764.6 (0.4%) | 0.003 |
| ...Florida | 2086 (2.2%) | 2220 (2.4%) | 0.013 | 4208.8<br>(2.3%) | 4313.8<br>(2.3%) | 0.001 |
| ...Georgia | 1053 (1.1%) | 684 (0.7%) | 0.039 | 1758.2<br>(1.0%) | 1774.3<br>(1.0%) | 0.002 |
| ...Hawaii | 146 (0.2%) | 114 (0.1%) | 0.008 | 263.1 (0.2%) | 268.7 (0.1%) | 0.000 |
| ...Iowa | 711 (0.8%) | 396 (0.4%) | 0.042 | 1115.9<br>(0.6%) | 1146.9<br>(0.6%) | 0.000 |
| ...Idaho/<br>Wyoming | 1381 (1.5%) | 1894 (2.1%) | 0.045 | 2968.8<br>(1.6%) | 3289.2<br>(1.8%) | 0.009 |
| ...Illinois | 9400 (9.9%) | 12,026<br>(13.0%) | 0.097 | 21,108.8<br>(11.7%) | 21,582.5<br>(11.6%) | 0.004 |
| ...Indiana | 497 (0.5%) | 826 (0.9%) | 0.044 | 1251.3<br>(0.7%) | 1322.1<br>(0.7%) | 0.002 |
| ...Kansas | 1432 (1.5%) | 2392 (2.6%) | 0.076 | 3493.9<br>(1.9%) | 3791.6<br>(2.0%) | 0.007 |
| ...Kentucky | 291 (0.3%) | 459 (0.5%) | 0.03 | 735.6 (0.4%) | 761.7 (0.4%) | 0.000 |
| ...Louisiana | 465 (0.5%) | 395 (0.4%) | 0.009 | 855.1 (0.5%) | 867.2 (0.5%) | 0.001 |
| ...Massachusett<br>s/Rhode Island | 337 (0.4%) | 354 (0.4%) | 0.004 | 682.9 (0.4%) | 696.9 (0.4%) | 0.001 |
| ...Maryland | 273 (0.3%) | 244 (0.3%) | 0.005 | 502.7 (0.3%) | 512.6 (0.3%) | 0.001 |
| ...Maine | 51 (0.1%) | 74 (0.1%) | 0.01 | 118.0 (0.1%) | 123.7 (0.1%) | 0.000 |
| ...Michigan | 9,859<br>(10.4%) | 10,890<br>(11.8%) | 0.043 | 19,454.1<br>(10.8%) | 20,620.2<br>(11.1%) | 0.009 |
| ...Minnesota | 152 (0.2%) | 163 (0.2%) | 0.004 | 313.9 (0.2%) | 321.7 (0.2%) | 0.000 |
| ...Missouri | 262 (0.3%) | 318 (0.3%) | 0.012 | 570.5 (0.3%) | 580.8 (0.3%) | 0.001 |
| ...Mississippi | 101 (0.1%) | 69 (0.1%) | 0.011 | 174.6 (0.1%) | 175.6 (0.1%) | 0.001 |
| ...Montana | 209 (0.2%) | 201 (0.2%) | 0.001 | 405.6 (0.2%) | 411.5 (0.2%) | 0.001 |
| ...North/South<br>Dakota | 34 (0.0%) | 55 (0.1%) | 0.011 | 80.3 (0.0%) | 86.9 (0.1%) | 0.001 |
| ...North Carolina | 330 (0.3%) | 476 (0.5%) | 0.025 | 760.5 (0.4%) | 806.2 (0.4%) | 0.002 |
| ...Nebraska | 288 (0.3%) | 696 (0.8%) | 0.062 | 862.7 (0.5%) | 981.5 (0.5%) | 0.007 |
| ...New<br>Hampshire | 33 (0.0%) | 39 (0.0%) | 0.004 | 74.8 (0.0%) | 73.7 (0.0%) | 0.001 |
| ...New Jersey | 5290 (5.6%) | 7834 (8.5%) | 0.113 | 12,507.0<br>(6.9%) | 13,120.7<br>(7.0%) | 0.004 |
| ...New Mexico | 1394 (1.5%) | 1066 (1.2%) | 0.028 | 2389.8<br>(1.3%) | 2437.1<br>(1.3%) | 0.002 |
| ...Nevada | 210 (0.2%) | 265 (0.3%) | 0.013 | 456.3 (0.3%) | 472.6 (0.3%) | 0.000 |
| ...New York | 3246 (3.4%) | 1828 (2.0%) | 0.09 | 5041.4<br>(2.8%) | 5024.6<br>(2.7%) | 0.006 |
| ...Ohio | 6178 (6.5%) | 7162 (7.8%) | 0.047 | 12,621.6<br>(7.0%) | 13,214.9<br>(7.1%) | 0.004 |
| ...Oklahoma | 727 (0.8%) | 997 (1.1%) | 0.032 | 1611.9<br>(0.9%) | 1726.4<br>(0.9%) | 0.003 |
| ...Oregon | 360 (0.4%) | 250 (0.3%) | 0.019 | 599.0 (0.3%) | 596.4 (0.3%) | 0.002 |
| ...Pennsylvania | 3454 (3.7%) | 2777 (3.0%) | 0.036 | 6256.8<br>(3.5%) | 6326.9<br>(3.4%) | 0.004 |
| ...South Carolina | 204 (0.2%) | 178 (0.2%) | 0.005 | 359.1 (0.2%) | 369.9 (0.2%) | 0.000 |
| ...Tennessee | 186 (0.2%) | 178 (0.2%) | 0.001 | 370.4 (0.2%) | 375.8 (0.2%) | 0.001 |
| ...Texas | 5740 (6.1%) | 6999 (7.6%) | 0.06 | 12,581.3<br>(7.0%) | 12,993.7<br>(7.0%) | 0.000 |
| ...Utah | 36 (0.0%) | 38 (0.0%) | 0.002 | 74.5 (0.0%) | 75.9 (0.0%) | 0.000 |
| ...Virginia | 3620 (3.8%) | 3242 (3.5%) | 0.017 | 6790.4<br>(3.8%) | 6954.9<br>(3.7%) | 0.002 |
| ...Vermont | 89 (0.1%) | 81 (0.1%) | 0.002 | 163.3 (0.1%) | 169.0 (0.1%) | 0.000 |
| ...Washington | 931 (1.0%) | 742 (0.8%) | 0.019 | 1633.0<br>(0.9%) | 1678.5<br>(0.9%) | 0.001 |
| ...Wisconsin | 5871 (6.2%) | 7281 (7.9%) | 0.065 | 12,037.7<br>(6.7%) | 12,862.1<br>(6.9%) | 0.009 |
| ...West Virginia | 38 (0.0%) | 31 (0.0%) | 0.003 | 65.9 (0.0%) | 66.7 (0.0%) | 0.000 |
| **Month and year**<br>**of index** |  |  |  |  |  |  |
| ...September 2021 | 634 (0.7%) | 8167 (8.8%) | 0.391 | 2831.4<br>(1.6%) | 8798.5<br>(4.7%) | 0.181 |
| ...October 2021 | 15,072<br>(15.9%) | 29,547<br>(32.0%) | 0.383 | 44,122.7<br>(24.4%) | 44,628.6<br>(23.9%) | 0.012 |
| ...November 2021 | 33,042<br>(34.9%) | 18,205<br>(19.7%) | 0.347 | 51,217.0<br>(28.4%) | 50,802.2<br>(27.2%) | 0.025 |
| ...December 2021 | 19,019<br>(20.1%) | 13,882<br>(15.0%) | 0.134 | 32,973.8<br>(18.3%) | 33,059.9<br>(17.7%) | 0.014 |
| ...January 2022 | 9597<br>(10.1%) | 7,690 (8.3%) | 0.063 | 17,318.5<br>(9.6%) | 17,322.0<br>(9.3%) | 0.011 |
| ...February 2022 | 2719 (2.9%) | 2410 (2.6%) | 0.016 | 5141.7<br>(2.9%) | 5132.6<br>(2.8%) | 0.006 |
| ...March 2022 | 1672 (1.8%) | 1459 (1.6%) | 0.015 | 3145.3<br>(1.7%) | 3134.7<br>(1.7%) | 0.005 |
| ...April 2022 | 5080 (5.4%) | 4007 (4.3%) | 0.048 | 9079.5<br>(5.0%) | 9033.1<br>(4.8%) | 0.009 |
| ...May 2022 | 3207 (3.4%) | 2751 (3.0%) | 0.023 | 5937.1<br>(3.3%) | 5907.0<br>(3.2%) | 0.007 |
| ...June 2022 | 1809 (1.9%) | 1628 (1.8%) | 0.011 | 3443.2<br>(1.9%) | 3423.6<br>(1.8%) | 0.005 |
| ...July 2022 | 2074 (2.2%) | 1975 (2.1%) | 0.004 | 4053.5<br>(2.2%) | 4041.2<br>(2.2%) | 0.005 |
| ...August 2022 | 662 (0.7%) | 656 (0.7%) | 0.001 | 1343.1<br>(0.7%) | 1334.8<br>(0.7%) | 0.003 |
| **Count of unique**<br>**NDC generic**<br>**names** |  |  | 0.054 |  |  | 0.006 |
| ...Mean (SD) | 8.13 (6.45) | 7.79 (6.34) |  | 7.99 (6.33) | 7.96 (6.49) |  |
| ...Median [IQR] | 7.00 [4.00,<br>11.00] | 7.00 [3.00,<br>11.00] |  | 7.00 [4.00,<br>11.00] | 7.00 [3.00,<br>11.00] |  |
| **Count of**<br>**hospitalization**<br>**events in 365-**<br>**day baseline** |  |  | 0.030 |  |  | 0.000 |
| **period** |  |  |  |  |  |  |
| ...Mean (SD) | 0.63 (3.80) | 0.76 (4.50) |  | 0.70 (4.53) | 0.70 (4.00) |  |
| ...Median [IQR] | 0.00 [0.00, 0.00] | 0.00 [0.00, 0.00] |  | 0.00 [0.00, 0.00] | 0.00 [0.00, 0.00] |  |
| **Count of outpatient events in 365-day baseline period** |  |  | 0.023 |  |  | 0.001 |
| ...Mean (SD) | 36.12 (70.33) | 37.79 (72.12) |  | 36.53 (71.06) | 36.56 (70.35) |  |
| ...Median [IQR] | 15.00 [8.00, 29.00] | 15.00 [8.00, 30.00] |  | 15.00 [8.00, 29.00] | 15.00 [8.00, 30.00] |  |
| **Comorbid conditions and scores** |  |  |  |  |  |  |
| **Charlson comorbidity score categories** |  |  |  |  |  |  |
| ...0 | 39,444 (41.7%) | 39,514 (42.8%) | 0.022 | 76,506.7 (42.4%) | 79,154.2 (42.4%) | 0.001 |
| ...1 | 18,145 (19.2%) | 17,681 (19.1%) | 0.001 | 34,483.1 (19.1%) | 35,663.0 (19.1%) | 0.000 |
| ...2+ | 36,998 (39.1%) | 35,182 (38.1%) | 0.021 | 69,617.1 (38.6%) | 71,801.0 (38.5%) | 0.002 |
| **Frailty score categories** |  |  |  |  |  |  |
| ...Robust (0-0.149) | 65,980 (69.8%) | 64,362 (69.7%) | 0.002 | 126,317.8 (69.9%) | 130,487.4 (69.9%) | 0.000 |
| ...Prefrail (0.15-0.249) | 25,077 (26.5%) | 23,865 (25.8%) | 0.015 | 47,138.0 (26.1%) | 48,616.2 (26.1%) | 0.001 |
| ...Mild frailty (0.25-0.349) | 3,039 (3.2%) | 3,573 (3.9%) | 0.035 | 6,135.3 (3.4%) | 6,460.2 (3.5%) | 0.004 |
| ...Moderate to severe frailty (≥0.35) | 491 (0.5%) | 577 (0.6%) | 0.014 | 1,015.8 (0.6%) | 1,054.4 (0.6%) | 0.000 |
| **Clinical conditions** |  |  |  |  |  |  |
| Alcohol use; n (%) | 2,909 (3.1%) | 2,685 (2.9%) | 0.010 | 5,455.8 (3.0%) | 5,619.7 (3.0%) | 0.001 |
| Arrhythmia; n (%) | 23,343<br>(24.7%) | 23,086<br>(25.0%) | 0.007 | 44,577.7<br>(24.7%) | 46,137.4<br>(24.7%) | 0.001 |
| Asthma; n (%) | 9,575<br>(10.1%) | 9,453<br>(10.2%) | 0.004 | 18,394.6<br>(10.2%) | 19,070.1<br>(10.2%) | 0.001 |
| Cancer; n (%) | 11,654<br>(12.3%) | 11,734<br>(12.7%) | 0.012 | 22,612.1<br>(12.5%) | 23,372.6<br>(12.5%) | 0.000 |
| Cardiovascular<br>disease; n (%) | 74,337<br>(78.6%) | 71,713<br>(77.6%) | 0.023 | 140,848.8<br>(78.0%) | 145,478.9<br>(78.0%) | 0.001 |
| Cerebrovascular<br>disease; n (%) | 11,855<br>(12.5%) | 11,630<br>(12.6%) | 0.002 | 22,607.3<br>(12.5%) | 23,335.8<br>(12.5%) | 0.000 |
| Coronary artery<br>disease; n (%) | 18,108<br>(19.1%) | 17,889<br>(19.4%) | 0.006 | 34,613.5<br>(19.2%) | 35,822.6<br>(19.2%) | 0.001 |
| Chronic kidney<br>disease; n (%) | 16,903<br>(17.9%) | 15,768<br>(17.1%) | 0.021 | 31,445.8<br>(17.4%) | 32,422.6<br>(17.4%) | 0.001 |
| Chronic lung<br>disease; n (%) | 19,605<br>(20.7%) | 18,654<br>(20.2%) | 0.013 | 37,081.6<br>(20.5%) | 38,284.9<br>(20.5%) | 0.000 |
| COPD; n (%) | 12,927<br>(13.7%) | 11,914<br>(12.9%) | 0.023 | 24,159.7<br>(13.4%) | 24,861.6<br>(13.3%) | 0.002 |
| Dementia; n (%) | 3,958 (4.2%) | 4,808 (5.2%) | 0.048 | 7,816.1<br>(4.3%) | 8,367.9<br>(4.5%) | 0.008 |
| Diabetes; n (%) | 29,535<br>(31.2%) | 27,571<br>(29.8%) | 0.030 | 55,275.7<br>(30.6%) | 56,880.1<br>(30.5%) | 0.003 |
| Down's<br>syndrome<sup>1</sup>; n (%) | 16 (0.0%) | 17 (0.0%) | 0.001 | 32.4 (0.0%) | 29.3 (0.0%) | 0.002 |
| Congestive heart<br>failure; n (%) | 9,695<br>(10.2%) | 9,680<br>(10.5%) | 0.008 | 18,564.1<br>(10.3%) | 19,176.1<br>(10.3%) | 0.000 |
| Hypertension; n<br>(%) | 67,001<br>(70.8%) | 64,366<br>(69.7%) | 0.025 | 126,655.8<br>(70.1%) | 130,743.6<br>(70.1%) | 0.002 |
| Irritable bowel<br>Syndrome; n (%) | 28,485<br>(30.1%) | 27,672<br>(30.0%) | 0.003 | 54,091.8<br>(30.0%) | 55,930.4<br>(30.0%) | 0.000 |
| Liver disease; n<br>(%) | 7,064 (7.5%) | 6,264 (6.8%) | 0.027 | 13,062.3<br>(7.2%) | 13,356.8<br>(7.2%) | 0.003 |
| Obesity; n (%) | 25,789<br>(27.3%) | 24,049<br>(26.0%) | 0.028 | 48,357.1<br>(26.8%) | 49,847.0<br>(26.7%) | 0.001 |
| Influenza or RSV;<br>n (%) | 542 (0.6%) | 619 (0.7%) | 0.012 | 1,101.7<br>(0.6%) | 1,153.2<br>(0.6%) | 0.001 |
| Psoriasis; n (%) | 832 (0.9%) | 837 (0.9%) | 0.003 | 1,632.2<br>(0.9%) | 1,687.3<br>(0.9%) | 0.000 |
| Psoriatic arthritis;<br>n (%) | 375 (0.4%) | 365 (0.4%) | 0.000 | 738.2 (0.4%) | 764.1 (0.4%) | 0.000 |
| Rheumatoid | 2243 (2.4%) | 1995 (2.2%) | 0.014 | 4155.4 | 4257.9 | 0.001 |
| arthritis; n (%) |  |  |  | (2.3%) | (2.3%) |  |
| Sickle cell disease or thalassemia; n (%) | 83 (0.1%) | 72 (0.1%) | 0.003 | 146.7 (0.1%) | 150.0 (0.1%) | 0.000 |
| History of tobacco use/smoking; n (%) | 23,008 (24.3%) | 22,110 (23.9%) | 0.009 | 43,646.6 (24.2%) | 45,081.2 (24.2%) | 0.000 |
| **Primary series<sup>1,2</sup>** |  |  |  |  |  |  |
| Homologous primary series mRNA-1273 (mRNA-1273) [second mRNA-1273 vaccine] [open and closed claims]; n (%) | 90,422 (95.6%) | 7,035 (7.6%) | 4.205 | 172,685.9 (95.6%) | 18,019.1 (9.7%) | 3.382 |
| Homologous primary series BNT162b2 (BNT-162b2) [second BNT162b2 vaccine] [open and closed claims]; n (%) | 4127 (4.4%) | 85,338 (92.4%) | 4.21 | 8,134.9 (4.5%) | 168,924.2 (90.5%) | 3.389 |
| Any heterologous primary series (any order); n (%) | 1162 (1.2%) | 216 (0.2%) | 0.126 | 2744.5 (1.5%) | 682.6 (0.4%) | 0.12 |

COPD, chronic obstructive pulmonary disease; IQR, interquartile range; RSV, respiratory syncytial virus; SD, standard deviation.

The greatest proportion of patients received mRNA-1273 and BNT162b2 in November 2021 and October 2021, respectively. Most participants in the mRNA-1273 and BNT162b2 groups had received a homologous primary series (95.6% and 92.4%, respectively). Baseline healthcare resource utilization and comorbid distributions were similar between the groups.

After applying inverse probability of treatment weighting, there was an effective sample size of 167,571 adults ≥65 years, of which 85,575 (51.1%) had received a third dose of mRNA-1273 and 81,996 (48.9%) had received a third dose of BNT162b2. Within this weighted population, the median age was 69 years (interquartile range, 66-74), 53.5% were female, 46% of participants were commercially insured, and 27% were insured by Medicare Advantage (Table 1). The most frequently occurring comorbidities included cardiovascular disease (78% in each exposure arm), hypertension (70.1% in each exposure arm), diabetes (30.6% and 30.5% for recipients of a third dose of mRNA-1273 and BNT162b2, respectively), and obesity (36.6% and 36.7% for recipients of a third dose of mRNA-1273 and BNT162b2, respectively). All covariates were considered well-balanced given ASDs of <0.10, with the exception of the month and year of cohort entry in the subgroup of adults that received a homologous primary series. Thus, models were adjusted for month and year of cohort entry in weighted analyses. All pre- and post-weighting baseline characteristics are presented in Table 1.

Results from the primary analysis (Table 2) indicated a lower rate of COVID-19 hospitalizations among those who received a third dose of mRNA-1273 (5.61 per 1000 PYs; 95% CI, 5.13-6.09) than those who received a third dose of BNT162b2 (7.06 per 1000 PYs; 95% CI, 6.54-7.57; HR, 0.82; 95% CI, 0.69-0.98) in the weighted population. The rate of medically attended COVID-19 was lower among adults with a third dose of mRNA-1273 (95.05 per 1000 PYs; 95% CI, 93.03-97.06) compared to BNT162b2 (106.55 per 1000 PYs; 95% CI, 104.53-108.57; HR, 0.93; 95% CI, 0.89-0.98).

**Table 2.**
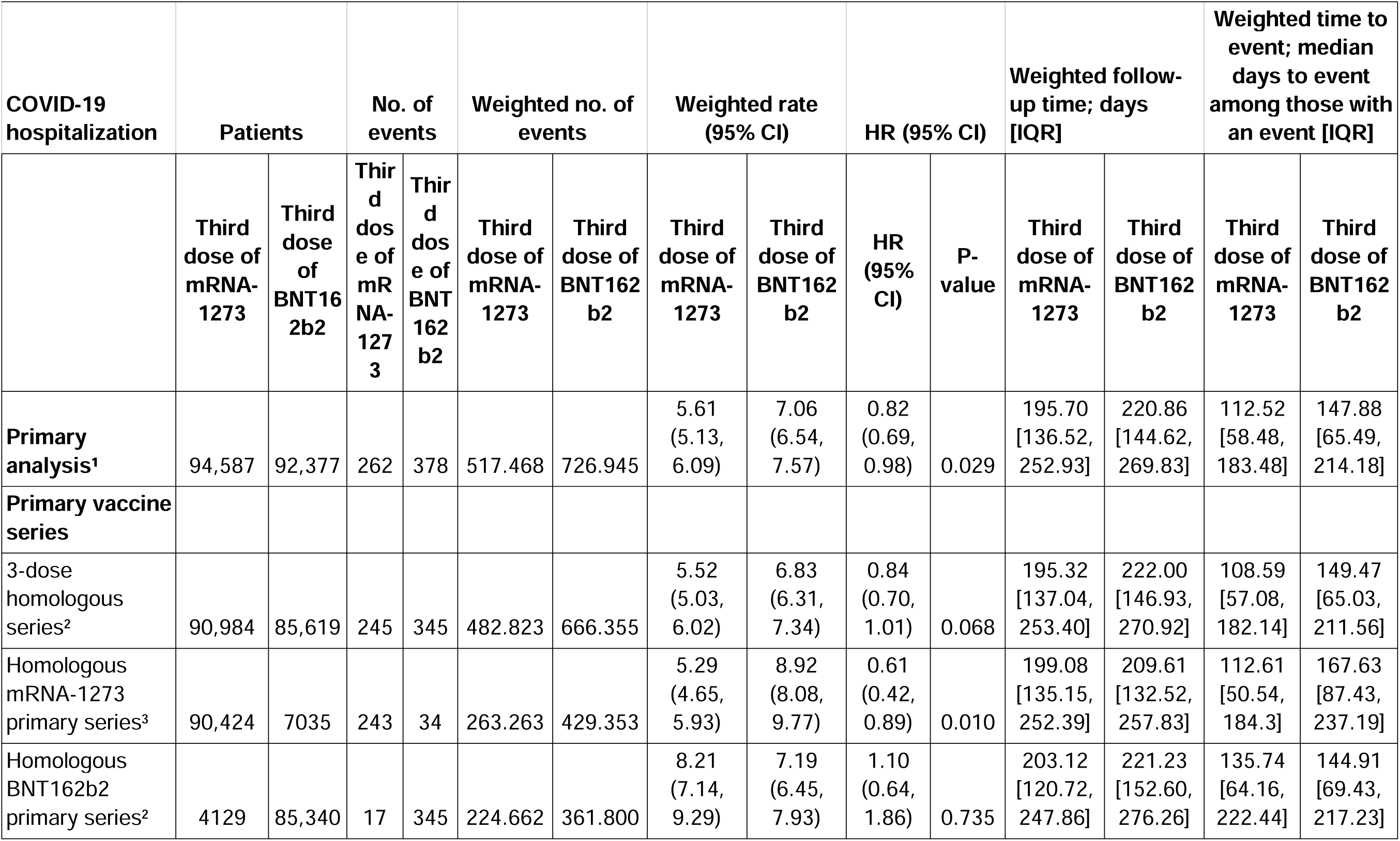
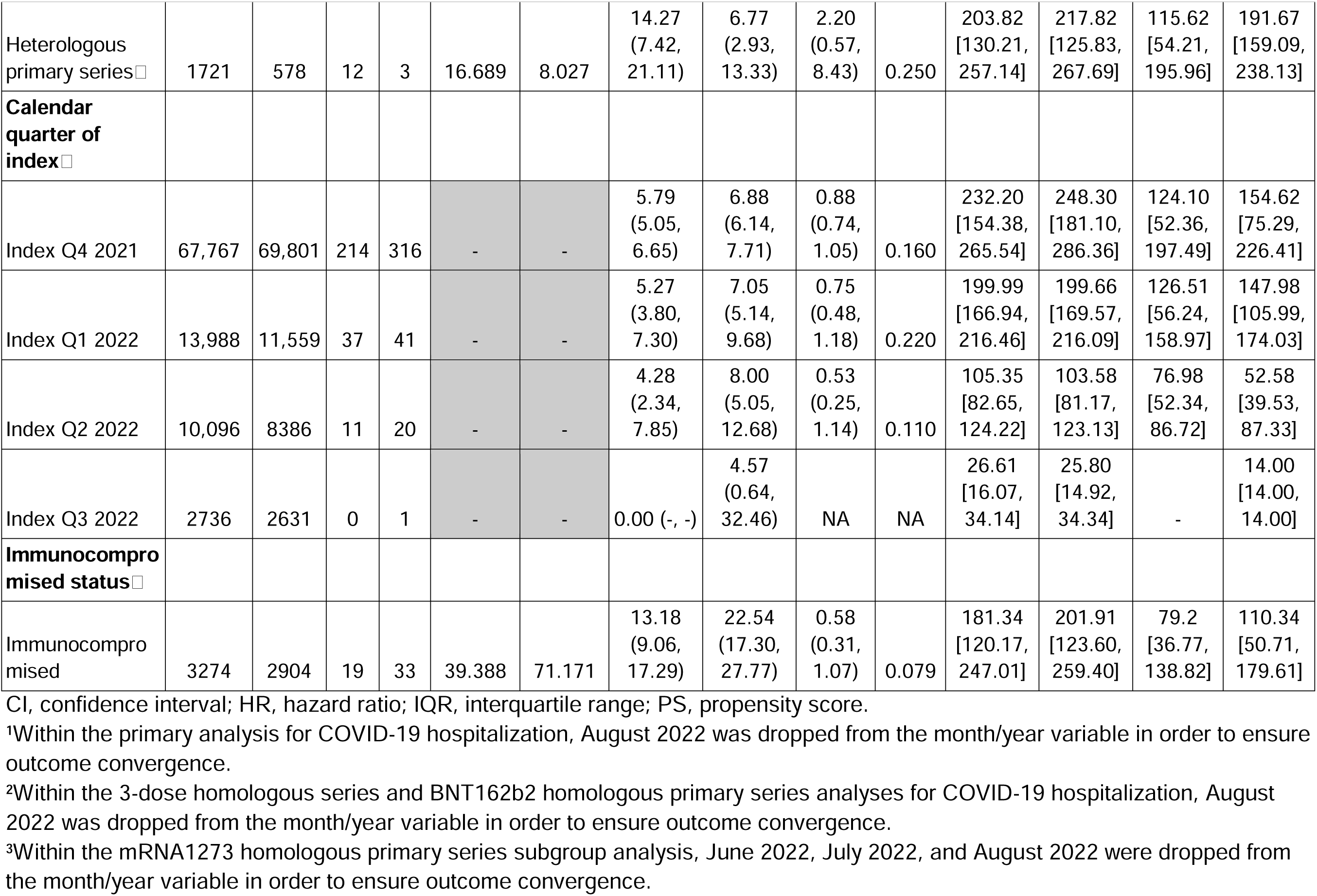
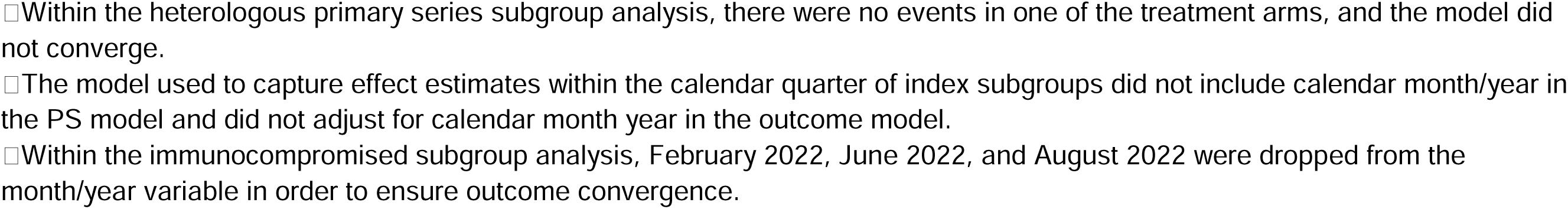
COVID-19 hospitalization effect estimates for the primary analysis and subgroups.

A subgroup of adults (n = 176,603) who received a 3-dose homologous series was included, among which 90,984 (51.5%) and 85,619 (48.5%) were vaccinated with mRNA-1273 and BNT162b2, respectively. Characteristics of the subgroup population were consistent with the overall population prior to weighting, with greater ASDs in the distribution of month and year of cohort entry between groups. Characteristics for the 3-dose homologous subgroup are available in Table S4.

The comparative analyses were repeated among a few pre-specified subgroups of interest. Within the subgroup of adults who received a 3-dose homologous series, COVID-19 hospitalization rates were directionally consistent, though not statistically significant (HR, 0.84; 95% CI, 0.70-1.01) (Table 2; Figure 3). Other subgroups, including homologous primary series for mRNA-1273, within the IC population, and by calendar quarter of cohort entry, were directionally consistent with the primary analysis, though not statistically significant. Subgroup analyses for individuals who received a homologous BNT162b2 primary series and a heterologous primary series were imprecise given the paucity of observed events.

**Figure 3.**
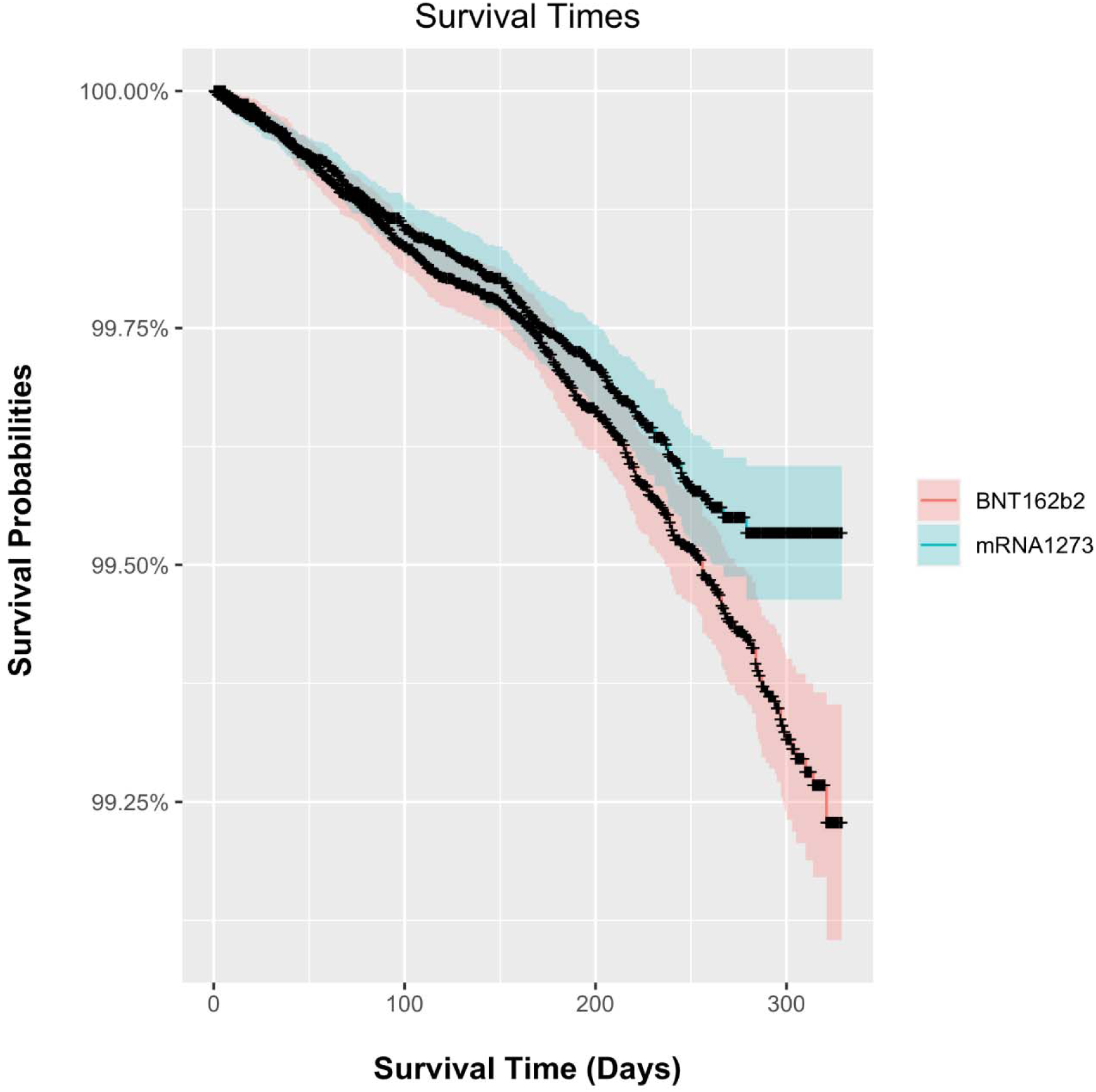
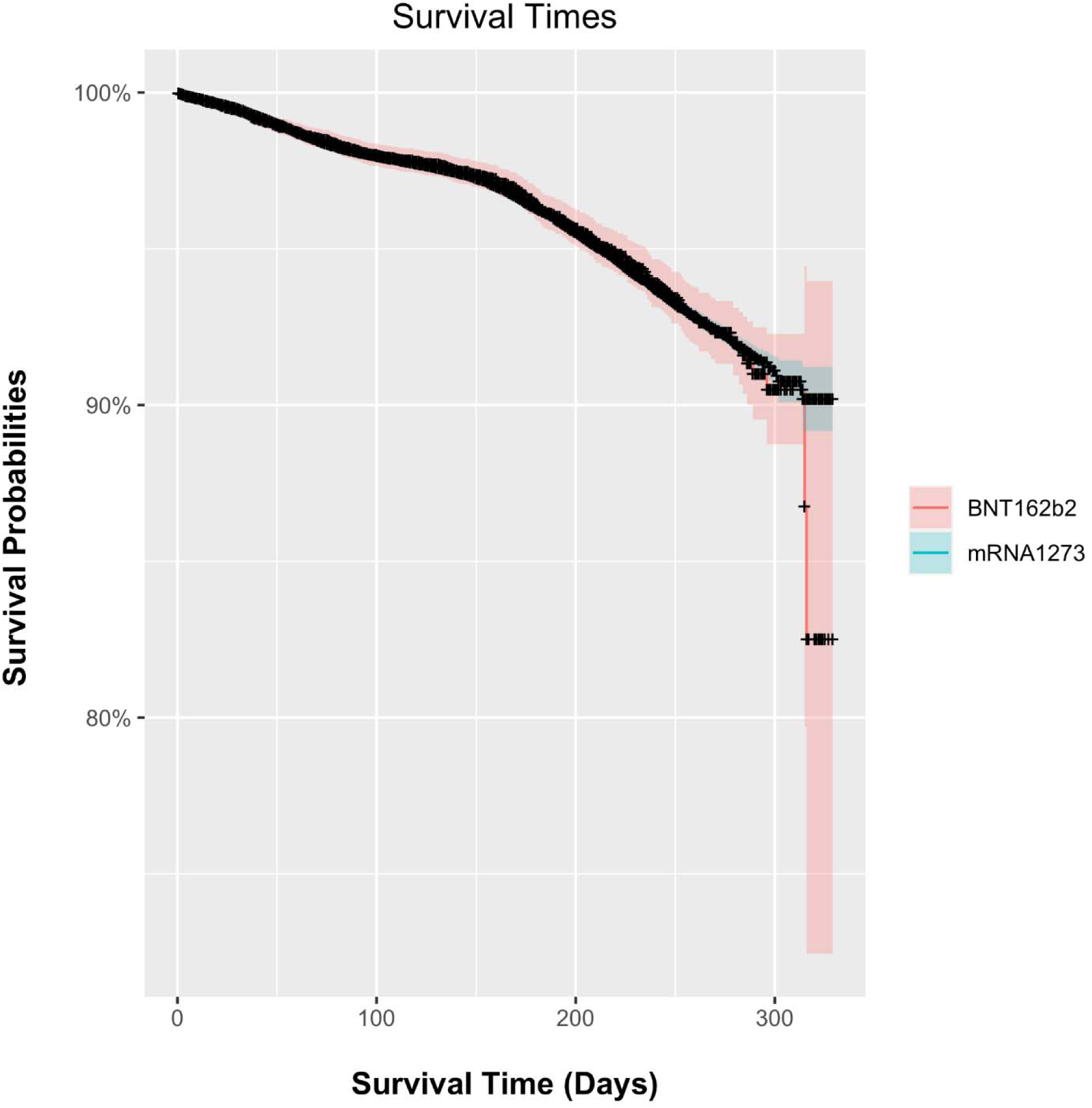
Comparative VE over time. Kaplan–Meier plots with 95% confidence intervals. A) COVID-19 hospitalization; B) medically attended COVID-19.

For the outcome of medically attended COVID-19, findings were consistent among the subgroup of adults who received a 3-dose homologous series (HR, 0.92; 95% CI, 0.88-0.97) as were those who received a heterologous primary series, within the IC population, and across calendar quarter of cohort entry, though these estimates were less precise given fewer observed events (Table 3). The subgroups of adults who received a homologous mRNA-1273 and BNT162b2 2-dose primary series were directionally different than the primary analysis, but also lacked statistical significance and precision given low event counts (Table 3).

**Table 3.**
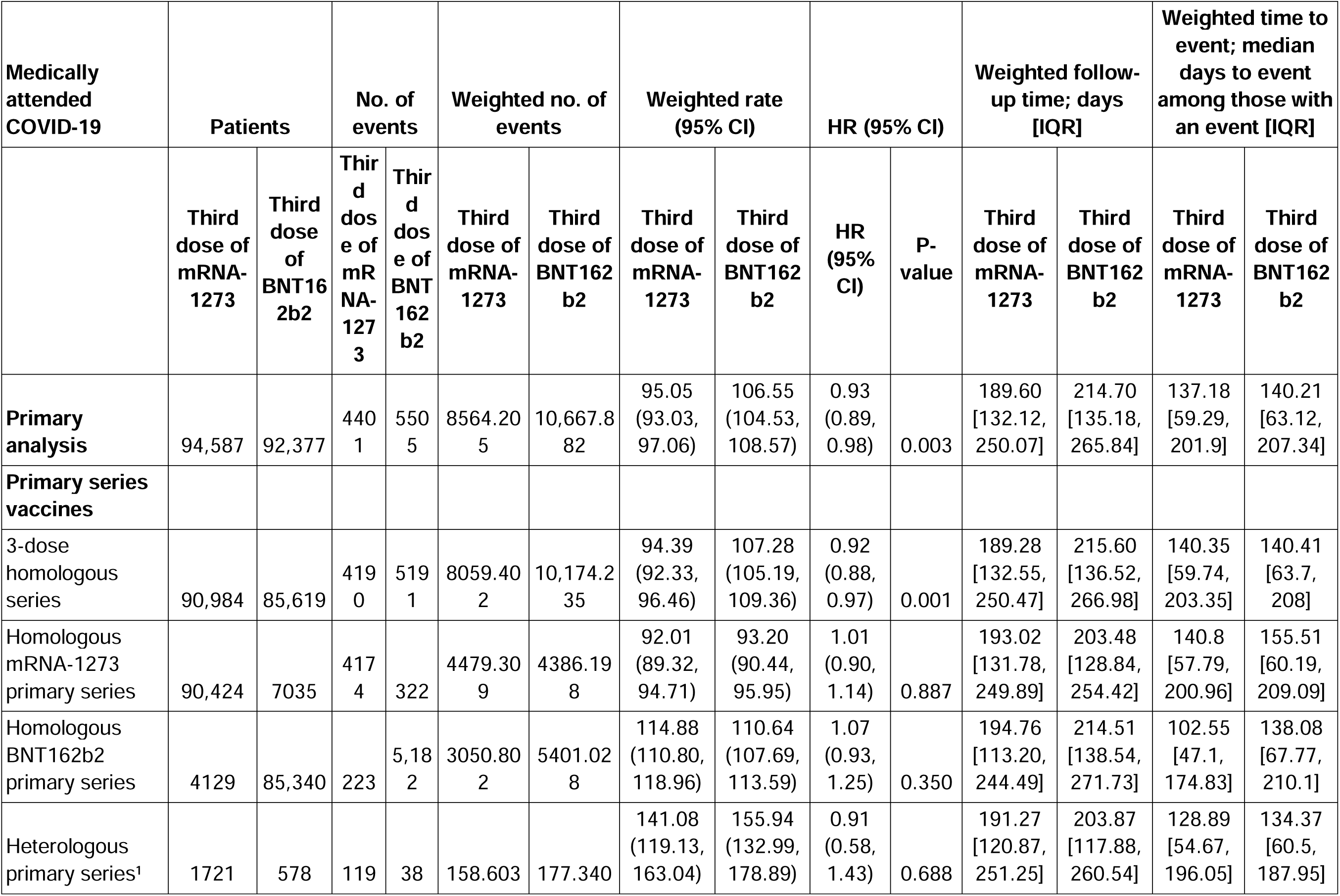
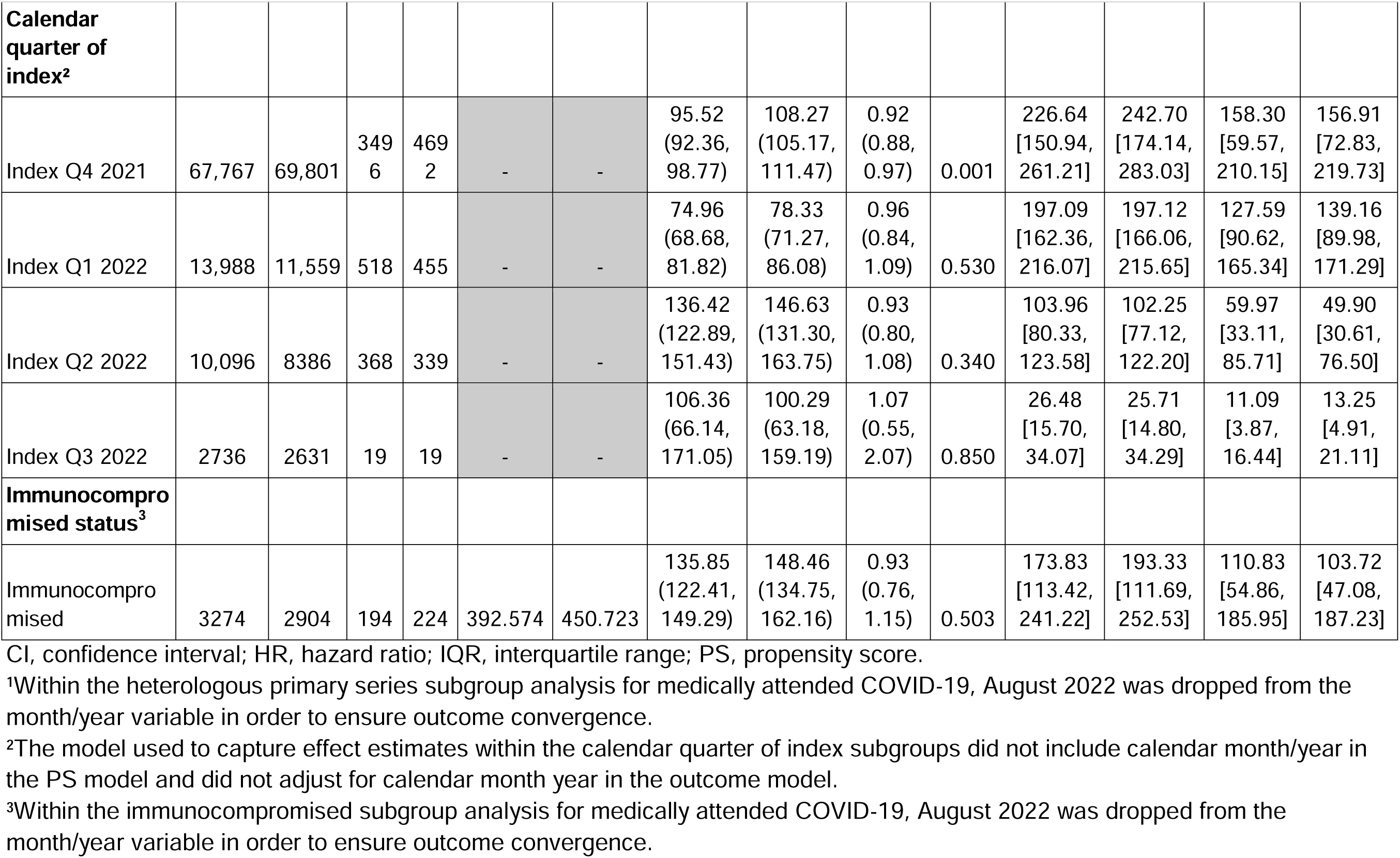
Medically attended COVID-19 effect estimates for primary analysis and subgroups.

Sensitivity analyses that were conducted using different outcome definitions led to consistent findings with the primary analyses (Tables S5 and S6).

## 4. Discussion

Older adult patients continue to be at risk for severe outcomes associated with COVID-19 infection. To date, there is limited information regarding the VE of booster doses of mRNA vaccines, particularly in the older adult population. Results of this observational study using a large US administrative claims database suggest that a third dose of mRNA-1273 is more effective in preventing COVID-19 hospitalization and medically attended COVID-19 than a third dose of BNT162b2 among insured older adults who completed a mRNA vaccination primary series.

Vaccine effectiveness has previously been shown to increase following a third dose of an mRNA vaccine [26]. A real-world study demonstrated a VE of 94% and 82% after a third dose of an mRNA vaccine during the delta and omicron variant infection periods, respectively [26]. The period over which patients were identified as having received a third dose of an mRNA COVID-19 vaccine in this study was at the tail-end of the delta variant surge in the United States in the later part of 2021 and overlapped with the surge of the omicron variants in 2022. VE and the results of the current study should be considered within the context of these variants. At the time of this study (as of March 29, 2022), additional booster doses were approved in the United States for both older adults and IC populations. Future work may consider the effectiveness of additional booster doses in this older adult population.

Our finding that a third dose of mRNA-1273 is more effective in preventing COVID-19 hospitalization and medically attended COVID-19 than BNT162b2 is consistent with real-world comparative effectiveness studies completed within boosted populations in both international and domestic data sources [10–14]. For example, among US veterans ≥18 years (median age, 70 years), adults who received a third dose of BNT162b2 compared with adults with a third dose of mRNA-1273 had a 16-week risk ratio of 1.15 (95% CI, 1.06-1.30) for documented SARS-CoV-2 infection, and 1.64 (95% CI, 1.27-2.79) for COVID-19 hospitalization [15]. Further, in a Spanish study of adults ≥40 years, leveraging Spain’s vaccination, laboratory, and national health system registries, adults who received a third dose of mRNA-1273 had a 13% lower risk of laboratory-confirmed COVID-19 infection than adults who received a third dose of BNT162b2 [11]. To the author’s knowledge, the current study represents the greatest sample of insured US adults ≥65 years examining the VE of a third dose of mRNA-1273 compared with BNT162b2, with findings that were consistent with the literature for both the primary analyses and within subgroups and sensitivity analyses.

In a subgroup of IC adults in this study, findings for both hospitalization of and medically attended COVID-19 were consistent with a prior analysis specific to the IC population, though effects observed in the current study were attenuated and not statistically significant. These differences could be attributed to the smaller sample size of IC adults aged ≥65 years, which was specific to a third dose of each manufacturer compared with a 2-dose series previously reported. Additionally, analysis of subgroups of individuals who received a homologous BNT162b2 2-dose primary series suggested a greater risk of each outcome among adults with a third dose of mRNA-1273 versus BNT162b2, though this was not statistically significant and lacked precision given the low proportion of events observed among the exposed coupled with a shorter follow-up time given differences in approval times between the vaccine manufacturers (203 and 223 days of follow-up). Similarly, a directionally different HR for COVID-19 hospitalization was observed among adults with a heterologous primary series, though there may not have been sufficient power to analyze results for this subgroup. Subgroup analyses by quarter year of index and among adults with a homologous mRNA-1273 primary series were consistent with primary analyses.

A strength of this head-to-head study is the utilization of a large claims database, which provided a sufficient sample size to detect differences between vaccine groups of less common outcomes (ie,COVID-19 hospitalization) among a population of older adults for whom the risk of breakthrough COVID-19 infection and hospitalization is higher than the general population. The sensitivity analyses incorporating open and closed claims for defining outcomes rather than closed claims alone produced results consistent with the primary analysis and provides support for incorporating open claims into capture of outcomes in future effectiveness studies.

Furthermore, inclusion of subgroups based on primary vaccine series and homologous 3-dose series provides contextualization for the impact of the primary series manufacturer on relative VE of a third dose and provides evidence that vaccination with a third dose of mRNA-1273 is more protective against COVID-19 hospitalization than BNT162b2 when a patient has received a 2-dose primary series of mRNA-1273.

This study has several limitations for consideration. Due to the nature of observational studies, there could be confounding bias in the absence of randomization. We included and adjusted for a wide range of measured potential confounders, including patient demographic, comorbidities, and healthcare utilizations. However, there could be residual confounding due to unmeasured variables, such lifestyle and social economic status. Given the claims-based approach to identifying the outcome of medically attended COVID-19 and COVID-19 hospitalization without accompanying laboratory confirmation, the potential to identify a false-negative diagnosis of COVID-19 and misclassify it as an outcome may exist. Claims from the Private Source 20 data stream do not indicate diagnosis position, which may limit the specificity of identifying COVID-19 hospitalizations. However, as a proxy for diagnosis position, the selected algorithm for COVID-19 hospitalization included a requirement for evidence of respiratory distress during the hospitalization during which a COVID-19 diagnosis was recorded. Although the algorithm in this study did not include a laboratory-confirmed diagnosis of COVID-19 infection or hospitalized COVID-19, any lack of sensitivity or specificity of the algorithm is likely to be non-differential across vaccine groups. Thus, any bias to the HR effect estimate is expected to be in the direction of null, and therefore, relative VE may be underestimated. The sensitivity analyses with alternative definitions for COVID-19 hospitalization, both the less restrictive capture using a COVID-19 diagnosis code in an inpatient setting and the more restrictive definition using a more limited set of respiratory distress diagnosis codes, provide assurance that the observed estimates for COVID-19 hospitalization are not the result of bias.

The outcome of medically attended COVID-19 identified via the U-code for COVID-19 diagnosis in the inpatient or outpatient setting has not been previously validated in the HealthVerity database. A published study in Veterans Assist data estimated the positive-predictive value of the ICD-10 code U07.1 to be 84.2% in all settings (inpatient, outpatient, and emergency department/urgent care) [27]. More recently, a validation of the COVID-19 intensive care unit code conducted among intensive care unit and step down patients reported a positive predictive value of 0.92 [28].

Use of administrative claims data presents challenges for capturing booster vaccines; in the current study, there was a risk of misclassifying the exposure of interest. The capture of a booster COVID-19 vaccine dose in claims data is limited to vaccine events that are processed through insurance claims records. Therefore, any vaccines that were administered in non-traditional clinical settings and not processed through insurance claims records (eg, a mass vaccination center) will not be observable in an administrative claims database. The result may be that a first observed vaccine dose is a patient’s third or fourth vaccine dose. This study mitigates the risk of misclassification by imposing a minimum and maximum time period for capturing a second dose derived from both data explorations and manufacturer guidelines.

While the capture of a third dose as a booster cannot be differentiated from the capture of a third primary series dose, the requirement for a third dose to occur ≥42 days following a primary series and after the Pfizer booster dose EUA increases the probability the dose identified as the index date is in actuality a booster vaccine dose.

Another limitation is the consideration of changing VE with respect to specific variants and the inability to confirm which variant caused individual COVID-19 infections and hospitalizations. The study period overlaps with the circulation of the delta and omicron variants in the United States. However, different regions within the United States had different variant peaks, which may have impacted local rates of infections and hospitalizations differently. While we were not able to measure and control specific variant impact, the potential bias introduced through variant-specific transmissibility is proxied through the inclusion of both state and calendar quarter of index in the propensity score model. In addition, this data source could not provide information for the Medicare fee-for-service population or uninsured elderly adults.

Importantly, as the COVID-19 vaccine landscape continues to evolve, many individuals have received additional vaccine doses beyond a third dose. Future work is needed to assess the comparative effectiveness of the updated XBB.1.5 mRNA COVID-19 vaccines.

In conclusion, results from this observational comparative VE database study provide evidence that among older adults, a third dose of mRNA-1273 is more effective in preventing breakthrough COVID-19 hospitalization and medically attended COVID-19 infection compared with a third dose of BNT162b2.

## Ethics committee approval statement

This study was fully approved by the WCG Institutional Review Board (#20231679)

## Funding

This work was supported by Moderna, Inc.

## Role of funding source

Authors employed by Moderna, Inc., were involved in the study design; in the collection, analysis, and interpretation of data; in the writing of the report; and in the decision to submit the paper for publication.

## Data sharing statement

Individual-level data reported in this study are not publicly shared. Upon request, and subject to review, Aetion, Inc., may provide the deidentified aggregate-level data that support the findings of this study. Deidentified data (including participant data as applicable) may be shared upon approval of an analysis proposal and a signed data access agreement.

## Declaration of interests

EB, LL, SSL, MG, MAM, TS, DBE, DM, and NV are employees of Moderna, Inc., and hold stock/stock options in the company. BK, CB, AT, and KEM are employees of Aetion, Inc., which has been contracted by Moderna, Inc., for the conduct of the present study. BK, CB, and KEM are stock option holders in Aetion, Inc.

## Acknowledgments

This study was funded by Moderna, Inc. Editorial assistance was provided by MEDiSTRAVA in accordance with Good Publication Practice (GPP3) guidelines, funded by Moderna, Inc., and under the direction of the authors.

## List of supplementary materials

Table S1 Vaccine codes.

Table S2. Baseline covariates.

Table S3. Immunocompromised status.

Table S4. Pre- and post-weighting characteristics for the 3-dose homologous series subgroup.

Table S5. COVID-19 hospitalization sensitivity analysis effect estimate.

Table S6. Medically attended COVID-19 sensitivity analysis effect estimate.

Supplementary file 1. Timing between first and second dose in a primary series and primary series to third dose

Supplementary file 2. Details on defining COVID-19 hospitalization

Supplementary file 3. Defining respiratory distress

## Notes

### Author Declarations

This study was fully approved by the WCG Institutional Review Board (#20231679)

