## Supplementary Materials for "Real-world comparative effectiveness of a third dose of mRNA-1273 versus BNT162b2 among adults aged ≥65 years in the United States"

Supplementary Tables

Table S1 Vaccine codes.

Table S2. Baseline covariates.

Table S3. Immunocompromised status.

Table S4. Pre- and post-weighting characteristics for the 3-dose homologous series subgroup.

Table S5. COVID-19 hospitalization sensitivity analysis effect estimate.

Table S6. Medically attended COVID-19 sensitivity analysis effect estimate.

**Table S1. Vaccine codes.**

| **Vaccine** | **Codes** |
| --- | --- |
| mRNA-1273 | **Primary vaccine codes**  CPT/HCPCS  0011A  0012A  91301  NDC  80777027310  80777027315  80777027398  80777027399  8077727315  8077727398  8077727399  8077727310  8077710011  8077701001  8077710099  80777010011  80777010099  CVX  207  **Booster vaccine codes**  CPT  91306  0064A  0094A  91309  91313  0134A  0013A  NDC  8077727505  80777027505  8077728205  80777028205  8077728299  80777028005  80777028099  80777028299  80777027599  8077727599  CVX  221  229 |
| BNT162b2 | **Primary vaccine codes**  CPT/HCPCS  91300  0001A  0002A  91305  0051A  0052A  NDC  59267100001  59267100002  59267100003  5926710001  5926710002  5926710003  59267102501  5926710251  00069202501  0006920251  00069202510  0069202501  0069202510  0069202525  00069202525  5926710252  59267102502  5926710253  59267102503  5926710254  59267102504  CVX  208  217  **Booster vaccine codes**  CPT/HCPCS  0004A  0054A  91312  0124A  0003A  0053A  NDC  59267030401  5926703041  5926703042  59267030402  59267140401  5926714041  5926714042  59267140402  CVX  300 |
| Janssen | CPT/HCPCS  91303  0031A  0034A  NDC  59676058005  59676058015  5967658005  5967658015  CVX  212 |
| AstraZeneca | CPT/HCPCS  91302  0021A  0022A  National Drug Code  0310122210  00310122210  00310122215  0310122215  CVX  210 |
| Novavax | CPT/HCPCS  0044A  91304  0041A  0042A  NDC  8063110001  80631100001  80631010001  80631010010  8063110010  CVX code  211 |

CPT, current procedural terminology; CVX, vaccine administered, Healthcare Common Procedure Coding System; NDC, national drug code.

**Table S2. Baseline covariates.**

| **Covariate category**  ***[assessment window]*** | **Specific patient characteristics** |
| --- | --- |
| Demographic variables  *[Index date]* | - Age (continuous) - Age (categorical)   - 65-74 years   - 75-84 years   - 85+ years - Sex (categorical: male, female, unknown/missing) - Insurance type (categorical: Commercial, Medicare, Medicaid) - State of residence |
| Healthcare resource utilization  *[Day -365 to Day -1]* | - Number of unique NDC generic name medications - Number of hospitalizations - Number of outpatient visits - Charlson comorbidity score categories   - 0   - 1   - 2+ - Frailty score categories   - Robust (0-<0.15)   - Prefrail (0.15-<0.25))   - Mild frailty (0.25-<0.35)   - Moderate to severe frailty (≥0.35) |
| Baseline clinical covariates  *[Start of all available data to Day -1]* | - Alcohol use - Arrhythmia - Asthma - Cancer - Cardiovascular disease - Cerebrovascular disease - Coronary artery disease - Chronic kidney disease - Chronic lung disease - COPD - Dementia - Diabetes (type II) - Down syndrome - Heart failure - Hypertension - Irritable bowel syndrome (IBS) - Liver disease - Obesity - Other respiratory viruses: influenza or RSV - Psoriasis - Psoriatic arthritis - Rheumatoid arthritis - Sickle cell disease or thalassemia - Tobacco use |
| Other  *[Index date]* | - Calendar month/year of index |

COPD, chronic obstructive pulmonary disease; NDC, national drug code; RSV, respiratory syncytial virus.

**Table S3. Immunocompromised status.**

| **Immunocompromised status sourced from Mues et al [20]** | | | | |
| --- | --- | --- | --- | --- |
| **Characteristic** | **Assessment window** | **Care settings** | **Diagnosis position** |  |
| Immunocompromised | Composite of other components | 1 IP claim or 2 OP claims | Any |  |
| Blood transplant | [-730, -1] | 1 IP claim or 2 OP claims | Any | ICD-10 procedure code  30230G4 Transfusion of Allogeneic Unspecified Bone Marrow into Peripheral Vein, Open Approach  30230X2 Transfusion of Allogeneic Related Cord Blood Stem Cells into Peripheral Vein, Open Approach  30230Y2 Transfusion of Allogeneic Related Hematopoietic Stem Cells into Peripheral Vein, Open Approach  30230Y3 Transfusion of Allogeneic Unrelated Hematopoietic Stem Cells into Peripheral Vein, Open Approach  30230Y4 Transfusion of Allogeneic Unspecified Hematopoietic Stem Cells into Peripheral Vein, Open Approach  30233AZ Transfusion of Embryonic Stem Cells into Peripheral Vein, Percutaneous Approach  30233G1 Transfusion of Nonautologous Bone Marrow into Peripheral Vein, Percutaneous Approach  30233Y3 Transfusion of Allogeneic Unrelated Hematopoietic Stem Cells into Peripheral Vein, Percutaneous Approach  30240G1 Transfusion of Nonautologous Bone Marrow into Central Vein, Open Approach  30240G2 Transfusion of Allogeneic Related Bone Marrow into Central Vein, Open Approach  30240G3 Transfusion of Allogeneic Unrelated Bone Marrow into Central Vein, Open Approach  30240X2 Transfusion of Allogeneic Related Cord Blood Stem Cells into Central Vein, Open Approach  30240X4 Transfusion of Allogeneic Unspecified Cord Blood Stem Cells into Central Vein, Open Approach  30240Y1 Transfusion of Nonautologous Hematopoietic Stem Cells into Central Vein, Open Approach  30243AZ Transfusion of Embryonic Stem Cells into Central Vein, Percutaneous Approach  30243G4 Transfusion of Allogeneic Unspecified Bone Marrow into Central Vein, Percutaneous Approach  30243Y2 Transfusion of Allogeneic Related Hematopoietic Stem Cells into Central Vein, Percutaneous Approach  30243Y4 Transfusion of Allogeneic Unspecified Hematopoietic Stem Cells into Central Vein, Percutaneous Approach  30250G1 Transfusion of Nonautologous Bone Marrow into Peripheral Artery, Open Approach  30253X1 Transfusion of Nonautologous Cord Blood Stem Cells into Peripheral Artery, Percutaneous Approach  30260G1 Transfusion of Nonautologous Bone Marrow into Central Artery, Open Approach  30260X1 Transfusion of Nonautologous Cord Blood Stem Cells into Central Artery, Open Approach  30263G1 Transfusion of Nonautologous Bone Marrow into Central Artery, Percutaneous Approach  30263Y1 Transfusion of Nonautologous Hematopoietic Stem Cells into Central Artery, Percutaneous Approach  30230AZ Transfusion of Embryonic Stem Cells into Peripheral Vein, Open Approach  30230G1 Transfusion of Nonautologous Bone Marrow into Peripheral Vein, Open Approach  30230G2 Transfusion of Allogeneic Related Bone Marrow into Peripheral Vein, Open Approach  30230G3 Transfusion of Allogeneic Unrelated Bone Marrow into Peripheral Vein, Open Approach  30230X1 Transfusion of Nonautologous Cord Blood Stem Cells into Peripheral Vein, Open Approach  30230X3 Transfusion of Allogeneic Unrelated Cord Blood Stem Cells into Peripheral Vein, Open Approach  30230X4 Transfusion of Allogeneic Unspecified Cord Blood Stem Cells into Peripheral Vein, Open Approach  30230Y1 Transfusion of Nonautologous Hematopoietic Stem Cells into Peripheral Vein, Open Approach  30233G2 Transfusion of Allogeneic Related Bone Marrow into Peripheral Vein, Percutaneous Approach  30233G3 Transfusion of Allogeneic Unrelated Bone Marrow into Peripheral Vein, Percutaneous Approach  30233G4 Transfusion of Allogeneic Unspecified Bone Marrow into Peripheral Vein, Percutaneous Approach  30233X1 Transfusion of Nonautologous Cord Blood Stem Cells into Peripheral Vein, Percutaneous Approach  30233X2 Transfusion of Allogeneic Related Cord Blood Stem Cells into Peripheral Vein, Percutaneous Approach  30233X3 Transfusion of Allogeneic Unrelated Cord Blood Stem Cells into Peripheral Vein, Percutaneous Approach  30233X4 Transfusion of Allogeneic Unspecified Cord Blood Stem Cells into Peripheral Vein, Percutaneous Approach  30233Y1 Transfusion of Nonautologous Hematopoietic Stem Cells into Peripheral Vein, Percutaneous Approach  30233Y2 Transfusion of Allogeneic Related Hematopoietic Stem Cells into Peripheral Vein, Percutaneous Approach  30233Y4 Transfusion of Allogeneic Unspecified Hematopoietic Stem Cells into Peripheral Vein, Percutaneous Approach  30240AZ Transfusion of Embryonic Stem Cells into Central Vein, Open Approach  30240G4 Transfusion of Allogeneic Unspecified Bone Marrow into Central Vein, Open Approach  30240X1 Transfusion of Nonautologous Cord Blood Stem Cells into Central Vein, Open Approach  30240X3 Transfusion of Allogeneic Unrelated Cord Blood Stem Cells into Central Vein, Open Approach  30240Y2 Transfusion of Allogeneic Related Hematopoietic Stem Cells into Central Vein, Open Approach  30240Y3 Transfusion of Allogeneic Unrelated Hematopoietic Stem Cells into Central Vein, Open Approach  30240Y4 Transfusion of Allogeneic Unspecified Hematopoietic Stem Cells into Central Vein, Open Approach  30243G1 Transfusion of Nonautologous Bone Marrow into Central Vein, Percutaneous Approach  30243G2 Transfusion of Allogeneic Related Bone Marrow into Central Vein, Percutaneous Approach  30243G3 Transfusion of Allogeneic Unrelated Bone Marrow into Central Vein, Percutaneous Approach  30243X1 Transfusion of Nonautologous Cord Blood Stem Cells into Central Vein, Percutaneous Approach  30243X2 Transfusion of Allogeneic Related Cord Blood Stem Cells into Central Vein, Percutaneous Approach  30243X3 Transfusion of Allogeneic Unrelated Cord Blood Stem Cells into Central Vein, Percutaneous Approach  30243X4 Transfusion of Allogeneic Unspecified Cord Blood Stem Cells into Central Vein, Percutaneous Approach  30243Y1 Transfusion of Nonautologous Hematopoietic Stem Cells into Central Vein, Percutaneous Approach  30243Y3 Transfusion of Allogeneic Unrelated Hematopoietic Stem Cells into Central Vein, Percutaneous Approach  30250X1 Transfusion of Nonautologous Cord Blood Stem Cells into Peripheral Artery, Open Approach  30250Y1 Transfusion of Nonautologous Hematopoietic Stem Cells into Peripheral Artery, Open Approach  30253G1 Transfusion of Nonautologous Bone Marrow into Peripheral Artery, Percutaneous Approach  30253Y1 Transfusion of Nonautologous Hematopoietic Stem Cells into Peripheral Artery, Percutaneous Approach  30260Y1 Transfusion of Nonautologous Hematopoietic Stem Cells into Central Artery, Open Approach  30263X1 Transfusion of Nonautologous Cord Blood Stem Cells into Central Artery, Percutaneous Approach  ICD-10 Diagnosis Code  D84.9 Immunodeficiency, unspecified  Z94.81 Bone marrow transplant status  D81.1 Severe combined immunodeficiency [SCID] with low T and B cell numbers  D84.81 Immunodeficiency due to conditions classified elsewhere  T86.00 Unspecified complication of bone marrow transplant  T86.09 Other complications of bone marrow transplant  Z48.290 Encounter for aftercare following bone marrow transplant |
| Stem cell transplant | [-730, -1] | 1 IP claim or 2 OP claims | Any | CPT/HCPCS  38240 Hematopoietic progenitor cell (HPC); allogeneic transplantation per donor  38242 Allogeneic lymphocyte infusions  38243 Hematopoietic progenitor cell (HPC); HPC boost  S2150 Bone marrow or blood  ICD-10 procedure code  30230G4 Transfusion of Allogeneic Unspecified Bone Marrow into Peripheral Vein, Open Approach  30230X2 Transfusion of Allogeneic Related Cord Blood Stem Cells into Peripheral Vein, Open Approach  30230Y2 Transfusion of Allogeneic Related Hematopoietic Stem Cells into Peripheral Vein, Open Approach  30230Y3 Transfusion of Allogeneic Unrelated Hematopoietic Stem Cells into Peripheral Vein, Open Approach  30230Y4 Transfusion of Allogeneic Unspecified Hematopoietic Stem Cells into Peripheral Vein, Open Approach  30233AZ Transfusion of Embryonic Stem Cells into Peripheral Vein, Percutaneous Approach  30233G1 Transfusion of Nonautologous Bone Marrow into Peripheral Vein, Percutaneous Approach  30233Y3 Transfusion of Allogeneic Unrelated Hematopoietic Stem Cells into Peripheral Vein, Percutaneous Approach  30240G1 Transfusion of Nonautologous Bone Marrow into Central Vein, Open Approach  30240G2 Transfusion of Allogeneic Related Bone Marrow into Central Vein, Open Approach  30240G3 Transfusion of Allogeneic Unrelated Bone Marrow into Central Vein, Open Approach  30240X2 Transfusion of Allogeneic Related Cord Blood Stem Cells into Central Vein, Open Approach  30240X4 Transfusion of Allogeneic Unspecified Cord Blood Stem Cells into Central Vein, Open Approach  30240Y0 Transfusion of Autologous Hematopoietic Stem Cells into Central Vein, Open Approach  30240Y1 Transfusion of Nonautologous Hematopoietic Stem Cells into Central Vein, Open Approach  30243AZ Transfusion of Embryonic Stem Cells into Central Vein, Percutaneous Approach  30243G4 Transfusion of Allogeneic Unspecified Bone Marrow into Central Vein, Percutaneous Approach  30243Y2 Transfusion of Allogeneic Related Hematopoietic Stem Cells into Central Vein, Percutaneous Approach  30243Y4 Transfusion of Allogeneic Unspecified Hematopoietic Stem Cells into Central Vein, Percutaneous Approach  30250G1 Transfusion of Nonautologous Bone Marrow into Peripheral Artery, Open Approach  30253X1 Transfusion of Nonautologous Cord Blood Stem Cells into Peripheral Artery, Percutaneous Approach  30260G1 Transfusion of Nonautologous Bone Marrow into Central Artery, Open Approach  30260X1 Transfusion of Nonautologous Cord Blood Stem Cells into Central Artery, Open Approach  30263G1 Transfusion of Nonautologous Bone Marrow into Central Artery, Percutaneous Approach  30263Y1 Transfusion of Nonautologous Hematopoietic Stem Cells into Central Artery, Percutaneous Approach  30230AZ Transfusion of Embryonic Stem Cells into Peripheral Vein, Open Approach  30230G1 Transfusion of Nonautologous Bone Marrow into Peripheral Vein, Open Approach  30230G2 Transfusion of Allogeneic Related Bone Marrow into Peripheral Vein, Open Approach  30230G3 Transfusion of Allogeneic Unrelated Bone Marrow into Peripheral Vein, Open Approach  30230X1 Transfusion of Nonautologous Cord Blood Stem Cells into Peripheral Vein, Open Approach  30230X3 Transfusion of Allogeneic Unrelated Cord Blood Stem Cells into Peripheral Vein, Open Approach  30230X4 Transfusion of Allogeneic Unspecified Cord Blood Stem Cells into Peripheral Vein, Open Approach  30230Y1 Transfusion of Nonautologous Hematopoietic Stem Cells into Peripheral Vein, Open Approach  30233G2 Transfusion of Allogeneic Related Bone Marrow into Peripheral Vein, Percutaneous Approach  30233G3 Transfusion of Allogeneic Unrelated Bone Marrow into Peripheral Vein, Percutaneous Approach  30233G4 Transfusion of Allogeneic Unspecified Bone Marrow into Peripheral Vein, Percutaneous Approach  30233X1 Transfusion of Nonautologous Cord Blood Stem Cells into Peripheral Vein, Percutaneous Approach  30233X2 Transfusion of Allogeneic Related Cord Blood Stem Cells into Peripheral Vein, Percutaneous Approach  30233X3 Transfusion of Allogeneic Unrelated Cord Blood Stem Cells into Peripheral Vein, Percutaneous Approach  30233X4 Transfusion of Allogeneic Unspecified Cord Blood Stem Cells into Peripheral Vein, Percutaneous Approach  30233Y1 Transfusion of Nonautologous Hematopoietic Stem Cells into Peripheral Vein, Percutaneous Approach  30233Y2 Transfusion of Allogeneic Related Hematopoietic Stem Cells into Peripheral Vein, Percutaneous Approach  30233Y4 Transfusion of Allogeneic Unspecified Hematopoietic Stem Cells into Peripheral Vein, Percutaneous Approach  30240AZ Transfusion of Embryonic Stem Cells into Central Vein, Open Approach  30240G4 Transfusion of Allogeneic Unspecified Bone Marrow into Central Vein, Open Approach  30240X1 Transfusion of Nonautologous Cord Blood Stem Cells into Central Vein, Open Approach  30240X3 Transfusion of Allogeneic Unrelated Cord Blood Stem Cells into Central Vein, Open Approach  30240Y2 Transfusion of Allogeneic Related Hematopoietic Stem Cells into Central Vein, Open Approach  30240Y3 Transfusion of Allogeneic Unrelated Hematopoietic Stem Cells into Central Vein, Open Approach  30240Y4 Transfusion of Allogeneic Unspecified Hematopoietic Stem Cells into Central Vein, Open Approach  30243G1 Transfusion of Nonautologous Bone Marrow into Central Vein, Percutaneous Approach  30243G2 Transfusion of Allogeneic Related Bone Marrow into Central Vein, Percutaneous Approach  30243G3 Transfusion of Allogeneic Unrelated Bone Marrow into Central Vein, Percutaneous Approach  30243X1 Transfusion of Nonautologous Cord Blood Stem Cells into Central Vein, Percutaneous Approach  30243X2 Transfusion of Allogeneic Related Cord Blood Stem Cells into Central Vein, Percutaneous Approach  30243X3 Transfusion of Allogeneic Unrelated Cord Blood Stem Cells into Central Vein, Percutaneous Approach  30243X4 Transfusion of Allogeneic Unspecified Cord Blood Stem Cells into Central Vein, Percutaneous Approach  30243Y1 Transfusion of Nonautologous Hematopoietic Stem Cells into Central Vein, Percutaneous Approach  30243Y3 Transfusion of Allogeneic Unrelated Hematopoietic Stem Cells into Central Vein, Percutaneous Approach  30250X1 Transfusion of Nonautologous Cord Blood Stem Cells into Peripheral Artery, Open Approach  30250Y1 Transfusion of Nonautologous Hematopoietic Stem Cells into Peripheral Artery, Open Approach  30253G1 Transfusion of Nonautologous Bone Marrow into Peripheral Artery, Percutaneous Approach  30253Y1 Transfusion of Nonautologous Hematopoietic Stem Cells into Peripheral Artery, Percutaneous Approach  30260Y1 Transfusion of Nonautologous Hematopoietic Stem Cells into Central Artery, Open Approach  30263X1 Transfusion of Nonautologous Cord Blood Stem Cells into Central Artery, Percutaneous Approach |
| Organ transplant with immunosuppressive therapy | [start of data, -1] | 1 IP claim or 2 OP claims | Any | ICD-10 diagnosis code  D84.9 Immunodeficiency, unspecified  T86.298 Other complications of heart transplant  T86.30 Unspecified complication of heart  T86.818 Other complications of lung transplant  T86.819 Unspecified complication of lung transplant  T86.898 Other complications of other transplanted tissue  T86.899 Unspecified complication of other transplanted tissue  Z48.24 Encounter for aftercare following lung transplant  Z48.280 Encounter for aftercare following heart  Z48.288 Encounter for aftercare following multiple organ transplant  Z94.2 Lung transplant status  D84.821 Immunodeficiency due to drugs  T86.10 Unspecified complication of kidney transplant  T86.19 Other complication of kidney transplant  T86.20 Unspecified complication of heart transplant  T86.39 Other complications of heart  T86.40 Unspecified complication of liver transplant  T86.49 Other complications of liver transplant  T86.858 Other complications of intestine transplant  T86.859 Unspecified complication of intestine transplant  Z48.21 Encounter for aftercare following heart transplant  Z48.22 Encounter for aftercare following kidney transplant  Z48.23 Encounter for aftercare following liver transplant  Z94.0 Kidney transplant status  Z94.1 Heart transplant status  Z94.3 Heart and lungs transplant status  Z94.4 Liver transplant status  Z94.82 Intestine transplant status  Z94.83 Pancreas transplant status  HCPCS/CPT  32852 Lung transplant, single; with cardiopulmonary bypass  33935 Heart  44135 Intestinal allotransplantation; from cadaver donor  44136 Intestinal allotransplantation; from living donor  47136 Liver allotransplantation; heterotopic, partial or whole, from cadaver or living donor, any age  50365 Renal allotransplantation, implantation of graft; with recipient nephrectomy  A4671 DISPOSABLE CYCLER SET USED WITH CYCLER DIALYSIS MACHINE, EACH  A4673 EXTENSION LINE WITH EASY LOCK CONNECTORS, USED WITH DIALYSIS  A4674 CHEMICALS/ANTISEPTICS SOLUTION USED TO CLEAN/STERILIZE DIALYSIS EQUIPMENT, PER 8 OZ  A4690 DIALYZER (ARTIFICIAL KIDNEYS), ALL TYPES, ALL SIZES, FOR HEMODIALYSIS, EACH  A4705 BICARBONATE DIALYSATE SOLUTION, EACH  A4706 BICARBONATE CONCENTRATE, SOLUTION, FOR HEMODIALYSIS, PER GALLON  A4707 BICARBONATE CONCENTRATE, POWDER, FOR HEMODIALYSIS, PER PACKET  A4708 ACETATE CONCENTRATE SOLUTION, FOR HEMODIALYSIS, PER GALLON  A4709 ACID CONCENTRATE, SOLUTION, FOR HEMODIALYSIS, PER GALLON  A4719 Y set tubing for peritoneal dialysis  A4721 DIALYSATE SOLUTION, ANY CONCENTRATION OF DEXTROSE, FLUID VOLUME GREATER THAN 999 CC BUT LESS THAN OR EQUAL TO 1999 CC, FOR PERITONEAL DIALYSIS  A4723 DIALYSATE SOLUTION, ANY CONCENTRATION OF DEXTROSE, FLUID VOLUME GREATER THAN 2999 CC BUT LESS THAN OR EQUAL TO 3999 CC, FOR PERITONEAL DIALYSIS  A4724 DIALYSATE SOLUTION, ANY CONCENTRATION OF DEXTROSE, FLUID VOLUME GREATER THAN 3999 CC BUT LESS THAN OR EQUAL TO 4999 CC, FOR PERITONEAL DIALYSIS  A4735 LOCAL/TOPICAL ANESTHETICS FOR DIALYSIS ONLY  A4737 INJECTABLE ANESTHETIC, FOR DIALYSIS, PER 10 ML  A4740 SHUNT ACCESSORY, FOR HEMODIALYSIS, ANY TYPE, EACH  A4750 BLOOD TUBING, ARTERIAL OR VENOUS, FOR HEMODIALYSIS, EACH  A4760 DIALYSATE SOLUTION TEST KIT, FOR PERITONEAL DIALYSIS, ANY TYPE, EACH  A4765 DIALYSATE CONCENTRATE, POWDER, ADDITIVE FOR PERITONEAL DIALYSIS, PER PACKET  A4820 HEMODIALYSIS KIT SUPPLIES  A4860 DISPOSABLE CATHETER TIPS FOR PERITONEAL DIALYSIS, PER 10  A4870 PLUMBING AND/OR ELECTRICAL WORK FOR HOME HEMODIALYSIS EQUIPMENT  A4890 CONTRACTS, REPAIR AND MAINTENANCE, FOR HEMODIALYSIS EQUIPMENT  A4900 CONTINUOUS AMBULATORY PERITONEAL DIALYSIS (CAPD) SUPPLY KIT  A4901 CONTINUOUS CYCLING PERITONEAL DIALYSIS (CCPD) SUPPLY KIT  A4911 DRAIN BAG/BOTTLE, FOR DIALYSIS, EACH  A4912 GOMCO DRAIN BOTTLE  A4913 MISCELLANEOUS DIALYSIS SUPPLIES, NOT OTHERWISE SPECIFIED  A4914 PREPARATION KITS  A4918 VENOUS PRESSURE CLAMP, FOR HEMODIALYSIS, EACH  E1510 Kidney, dialysate delivery system kidney machine, pump recirculating, air removal system, flowrate meter, power off, heater and temperature control with alarm, IV poles, pressure gauge, concentrate container  E1530 AIR BUBBLE DETECTOR FOR HEMODIALYSIS, EACH, REPLACEMENT  E1550 BATH CONDUCTIVITY METER FOR HEMODIALYSIS, EACH  E1590 HEMODIALYSIS MACHINE  E1594 CYCLER DIALYSIS MACHINE FOR PERITONEAL DIALYSIS  E1600 DELIVERY AND/OR INSTALLATION CHARGES FOR HEMODIALYSIS EQUIPMENT  E1615 DEIONIZER WATER PURIFICATION SYSTEM, FOR HEMODIALYSIS  E1635 COMPACT (PORTABLE) TRAVEL HEMODIALYZER SYSTEM  S2053 TRANSPLANTATION OF SMALL INTESTINE AND LIVER ALLOGRAFTS  S2054 TRANSPLANTATION OF MULTIVISCERAL ORGANS  S2142 CORD BLOOD  32851 Lung transplant, single; without cardiopulmonary bypass  32853 Lung transplant, double (bilateral sequential or en bloc); without cardiopulmonary bypass  32854 Lung transplant, double (bilateral sequential or en bloc); with cardiopulmonary bypass  33945 Heart transplant, with or without recipient cardiectomy  47135 Liver allotransplantation, orthotopic, partial or whole, from cadaver or living donor, any age  48554 Transplantation of pancreatic allograft  50360 Renal allotransplantation, implantation of graft; without recipient nephrectomy  50370 Removal of transplanted renal allograft  A4653 PERITONEAL DIALYSIS CATHETER ANCHORING DEVICE, BELT, EACH  A4672 DRAINAGE EXTENSION LINE, STERILE, FOR DIALYSIS, EACH  A4680 ACTIVATED CARBON FILTER FOR HEMODIALYSIS, EACH  A4700 STANDARD DIALYSATE SOLUTION, EACH  A4712 WATER, STERILE, FOR INJECTION, PER 10 ML  A4714 TREATED WATER (DEIONIZED, DISTILLED, OR REVERSE OSMOSIS) FOR PERITONEAL DIALYSIS, PER GALLON  A4720 DIALYSATE SOLUTION, ANY CONCENTRATION OF DEXTROSE, FLUID VOLUME GREATER THAN 249 CC, BUT LESS THAN OR EQUAL TO 999 CC, FOR PERITONEAL DIALYSIS  A4722 DIALYSATE SOLUTION, ANY CONCENTRATION OF DEXTROSE, FLUID VOLUME GREATER THAN 1999 CC BUT LESS THAN OR EQUAL TO 2999 CC, FOR PERITONEAL DIALYSIS  A4725 DIALYSATE SOLUTION, ANY CONCENTRATION OF DEXTROSE, FLUID VOLUME GREATER THAN 4999 CC BUT LESS THAN OR EQUAL TO 5999 CC, FOR PERITONEAL DIALYSIS  A4726 DIALYSATE SOLUTION, ANY CONCENTRATION OF DEXTROSE, FLUID VOLUME GREATER THAN 5999 CC, FOR PERITONEAL DIALYSIS  A4728 Dialysate solution, nondextrose containing, 500 ml  A4730 FISTULA CANNULATION SET FOR HEMODIALYSIS, EACH  A4736 Topical anesthetic, for dialysis, per g  A4755 BLOOD TUBING, ARTERIAL AND VENOUS COMBINED, FOR HEMODIALYSIS, EACH  A4766 DIALYSATE CONCENTRATE, SOLUTION, ADDITIVE FOR PERITONEAL DIALYSIS, PER 10 ML  A4802 PROTAMINE SULFATE, FOR HEMODIALYSIS, PER 50 MG  A4850 HEMOSTATS WITH RUBBER TIPS FOR DIALYSIS  A4880 STORAGE TANKS UTILIZED IN CONNECTION WITH WATER PURIFICATION SYSTEM, REPLACEMENT TANKS FOR DIALYSIS  A4905 INTERMITTENT PERITONEAL DIALYSIS (IPD) SUPPLY KIT  A4910 NON  E1500 CENTRIFUGE, FOR DIALYSIS  E1520 HEPARIN INFUSION PUMP FOR HEMODIALYSIS  E1540 PRESSURE ALARM FOR HEMODIALYSIS, EACH, REPLACEMENT  E1560 BLOOD LEAK DETECTOR FOR HEMODIALYSIS, EACH, REPLACEMENT  E1570 ADJUSTABLE CHAIR, FOR ESRD PATIENTS  E1575 TRANSDUCER PROTECTORS/FLUID BARRIERS, FOR HEMODIALYSIS, ANY SIZE, PER 10  E1580 UNIPUNCTURE CONTROL SYSTEM FOR HEMODIALYSIS  E1592 AUTOMATIC INTERMITTENT PERITONEAL DIALYSIS SYSTEM  E1610 REVERSE OSMOSIS WATER PURIFICATION SYSTEM, FOR HEMODIALYSIS  E1620 BLOOD PUMP FOR HEMODIALYSIS, REPLACEMENT  E1625 WATER SOFTENING SYSTEM, FOR HEMODIALYSIS  E1630 RECIPROCATING PERITONEAL DIALYSIS SYSTEM  E1632 WEARABLE ARTIFICIAL KIDNEY, EACH  E1634 PERITONEAL DIALYSIS CLAMPS, EACH  E1636 SORBENT CARTRIDGES, FOR HEMODIALYSIS, PER 10  S2060 LOBAR LUNG TRANSPLANTATION  S2065 SIMULTANEOUS PANCREAS KIDNEY TRANSPLANTATION  S2152 Solid organ(s), complete or segmental, single organ or combination of organs; deceased or living donor(s), procurement, transplantation, and related complications; including drugs; supplies; hospitalization with outpatient follow  ICD-10 procedure code  02YA0Z2 Transplantation of Heart, Zooplastic, Open Approach  07YM0Z0 Transplantation of Thymus, Allogeneic, Open Approach  07YM0Z1 Transplantation of Thymus, Syngeneic, Open Approach  07YP0Z0 Transplantation of Spleen, Allogeneic, Open Approach  07YP0Z1 Transplantation of Spleen, Syngeneic, Open Approach  07YP0Z2 Transplantation of Spleen, Zooplastic, Open Approach  0BYC0Z0 Transplantation of Right Upper Lung Lobe, Allogeneic, Open Approach  0BYC0Z1 Transplantation of Right Upper Lung Lobe, Syngeneic, Open Approach  0BYD0Z0 Transplantation of Right Middle Lung Lobe, Allogeneic, Open Approach  0BYD0Z1 Transplantation of Right Middle Lung Lobe, Syngeneic, Open Approach  0BYD0Z2 Transplantation of Right Middle Lung Lobe, Zooplastic, Open Approach  0BYF0Z0 Transplantation of Right Lower Lung Lobe, Allogeneic, Open Approach  0BYF0Z1 Transplantation of Right Lower Lung Lobe, Syngeneic, Open Approach  0BYF0Z2 Transplantation of Right Lower Lung Lobe, Zooplastic, Open Approach  0BYG0Z1 Transplantation of Left Upper Lung Lobe, Syngeneic, Open Approach  0BYH0Z0 Transplantation of Lung Lingula, Allogeneic, Open Approach  0BYL0Z0 Transplantation of Left Lung, Allogeneic, Open Approach  0BYM0Z1 Transplantation of Bilateral Lungs, Syngeneic, Open Approach  0DY80Z0 Transplantation of Small Intestine, Allogeneic, Open Approach  0DYE0Z0 Transplantation of Large Intestine, Allogeneic, Open Approach  0DYE0Z1 Transplantation of Large Intestine, Syngeneic, Open Approach  0FYG0Z1 Transplantation of Pancreas, Syngeneic, Open Approach  0TY00Z2 Transplantation of Right Kidney, Zooplastic, Open Approach  0TY10Z1 Transplantation of Left Kidney, Syngeneic, Open Approach  5A1D00Z Performance of Urinary Filtration, Single  5A1D70Z Performance of Urinary Filtration, Intermittent, Less than 6 Hours Per Day  5A1D80Z Performance of Urinary Filtration, Prolonged Intermittent, 6  5A1D90Z Performance of Urinary Filtration, Continuous, Greater than 18 hours Per Day  BT29ZZZ Computerized Tomography (CT Scan) of Kidney Transplant  BT39Y0Z Magnetic Resonance Imaging (MRI) of Kidney Transplant using Other Contrast, Unenhanced and Enhanced  02YA0Z0 Transplantation of Heart, Allogeneic, Open Approach  02YA0Z1 Transplantation of Heart, Syngeneic, Open Approach  07YM0Z2 Transplantation of Thymus, Zooplastic, Open Approach  0BYC0Z2 Transplantation of Right Upper Lung Lobe, Zooplastic, Open Approach  0BYG0Z0 Transplantation of Left Upper Lung Lobe, Allogeneic, Open Approach  0BYG0Z2 Transplantation of Left Upper Lung Lobe, Zooplastic, Open Approach  0BYH0Z1 Transplantation of Lung Lingula, Syngeneic, Open Approach  0BYH0Z2 Transplantation of Lung Lingula, Zooplastic, Open Approach  0BYJ0Z0 Transplantation of Left Lower Lung Lobe, Allogeneic, Open Approach  0BYJ0Z1 Transplantation of Left Lower Lung Lobe, Syngeneic, Open Approach  0BYJ0Z2 Transplantation of Left Lower Lung Lobe, Zooplastic, Open Approach  0BYK0Z0 Transplantation of Right Lung, Allogeneic, Open Approach  0BYK0Z1 Transplantation of Right Lung, Syngeneic, Open Approach  0BYK0Z2 Transplantation of Right Lung, Zooplastic, Open Approach  0BYL0Z1 Transplantation of Left Lung, Syngeneic, Open Approach  0BYL0Z2 Transplantation of Left Lung, Zooplastic, Open Approach  0BYM0Z0 Transplantation of Bilateral Lungs, Allogeneic, Open Approach  0BYM0Z2 Transplantation of Bilateral Lungs, Zooplastic, Open Approach  0DY50Z0 Transplantation of Esophagus, Allogeneic, Open Approach  0DY50Z1 Transplantation of Esophagus, Syngeneic, Open Approach  0DY50Z2 Transplantation of Esophagus, Zooplastic, Open Approach  0DY60Z0 Transplantation of Stomach, Allogeneic, Open Approach  0DY60Z1 Transplantation of Stomach, Syngeneic, Open Approach  0DY60Z2 Transplantation of Stomach, Zooplastic, Open Approach  0DY80Z1 Transplantation of Small Intestine, Syngeneic, Open Approach  0DY80Z2 Transplantation of Small Intestine, Zooplastic, Open Approach  0DYE0Z2 Transplantation of Large Intestine, Zooplastic, Open Approach  0FY00Z0 Transplantation of Liver, Allogeneic, Open Approach  0FY00Z1 Transplantation of Liver, Syngeneic, Open Approach  0FY00Z2 Transplantation of Liver, Zooplastic, Open Approach  0FYG0Z0 Transplantation of Pancreas, Allogeneic, Open Approach  0FYG0Z2 Transplantation of Pancreas, Zooplastic, Open Approach  0TY00Z0 Transplantation of Right Kidney, Allogeneic, Open Approach  0TY00Z1 Transplantation of Right Kidney, Syngeneic, Open Approach  0TY10Z0 Transplantation of Left Kidney, Allogeneic, Open Approach  0TY10Z2 Transplantation of Left Kidney, Zooplastic, Open Approach  3E1M39Z Irrigation of Peritoneal Cavity using Dialysate, Percutaneous Approach  BT2900Z Computerized Tomography (CT Scan) of Kidney Transplant using High Osmolar Contrast, Unenhanced and Enhanced  BT290ZZ Computerized Tomography (CT Scan) of Kidney Transplant using High Osmolar Contrast  BT2910Z Computerized Tomography (CT Scan) of Kidney Transplant using Low Osmolar Contrast, Unenhanced and Enhanced  BT291ZZ Computerized Tomography (CT Scan) of Kidney Transplant using Low Osmolar Contrast  BT29Y0Z Computerized Tomography (CT Scan) of Kidney Transplant using Other Contrast, Unenhanced and Enhanced  BT29YZZ Computerized Tomography (CT Scan) of Kidney Transplant using Other Contrast  BT39YZZ Magnetic Resonance Imaging (MRI) of Kidney Transplant using Other Contrast  BT39ZZZ Magnetic Resonance Imaging (MRI) of Kidney Transplant  BT49ZZZ Ultrasonography of Kidney Transplant  Diagnosis Related Group (Standard)  103  302  480  495  Immunosuppressive therapy  Generic name  BARICITINIB  BASILIXIMAB  BRODALUMAB  CANAKINUMAB/PF  DECITABINE/CEDAZURIDINE  ECULIZUMAB  FLUOROURACIL/ADHESIVE BANDAGE  GUSELKUMAB  INFLIXIMAB  INFLIXIMAB-ABDA  INFLIXIMAB-DYYB  NELARABINE  OCRELIZUMAB  PEMETREXED DISODIUM  PIRFENIDONE  PRALATREXATE  RAVULIZUMAB-CWVZ  ANAKINRA  BELATACEPT  CARMUSTINE IN POLIFEPROSAN 20  CERTOLIZUMAB PEGOL  CYCLOSPORINE/CHONDROITIN SULFATE A SODIUM  DACLIZUMAB  DAUNORUBICIN/CYTARABINE LIPOSOMAL  FINGOLIMOD HCL  GEMCITABINE HCL IN 0.9 % SODIUM CHLORIDE  GOLIMUMAB  INEBILIZUMAB-CDON  INFLIXIMAB-AXXQ  SATRALIZUMAB-MWGE  SECUKINUMAB  STREPTOZOCIN  TACROLIMUS IN VEHICLE BASE NO.238  TEPROTUMUMAB-TRBW  VEDOLIZUMAB  ABATACEPT  ABATACEPT/MALTOSE  ALEFACEPT  ALEMTUZUMAB  CYTARABINE LIPOSOME/PF  RISANKIZUMAB-RZAA  SARILUMAB  TACROLIMUS, MICRONIZED  TACROLIMUS/NIACINAMIDE  TERIFLUNOMIDE  USTEKINUMAB  AZATHIOPRINE SODIUM  DIROXIMEL FUMARATE  EFALIZUMAB  MELPHALAN HCL/BETADEX SULFOBUTYL ETHER SODIUM  MUROMONAB-CD3  MYCOPHENOLATE MOFETIL HCL  NATALIZUMAB  OZANIMOD HYDROCHLORIDE  RILONACEPT  TACROLIMUS/HYALURONATE SODIUM/NIACINAMIDE  TOCILIZUMAB  UPADACITINIB  APREMILAST  BELIMUMAB  BENDAMUSTINE HCL  FLOXURIDINE  SIPONIMOD  TEMSIROLIMUS  TILDRAKIZUMAB-ASMN  IXEKIZUMAB  POMALIDOMIDE  EMAPALUMAB-LZSG  OFATUMUMAB  TACROLIMUS ANHYDROUS  IFOSFAMIDE/MESNA  SILTUXIMAB  TOFACITINIB CITRATE  CLOFARABINE  LENALIDOMIDE  LOMUSTINE  CARMUSTINE  MELPHALAN  THALIDOMIDE  FLUDARABINE PHOSPHATE  CYTARABINE/PF  ETANERCEPT  AZACITIDINE  CLADRIBINE  THIOTEPA  CHLORAMBUCIL  ADALIMUMAB  IFOSFAMIDE  METHOTREXATE  MELPHALAN HCL  CYTARABINE  DECITABINE  DACARBAZINE  MYCOPHENOLATE SODIUM  DIMETHYL FUMARATE  MERCAPTOPURINE  BUSULFAN  CYCLOSPORINE, MODIFIED  LEFLUNOMIDE  SIROLIMUS  METHOTREXATE/PF  EVEROLIMUS  CAPECITABINE  CYCLOSPORINE  AZATHIOPRINE  GEMCITABINE HCL  METHOTREXATE SODIUM/PF  MYCOPHENOLATE MOFETIL  CYCLOPHOSPHAMIDE  METHOTREXATE SODIUM  FLUOROURACIL  TACROLIMUS  TEMOZOLOMIDE  HCPCS/CPT  80158 Cyclosporine  80169 Everolimus  80195 Sirolimus  80197 Tacrolimus  C9006 INJECTION, TACROLIMUS, PER 5 MG (1 AMP)  C9020 SIROLIMUS TABLET, 1 MG  C9026 Injection, vedolizumab, 1 mg  C9106 SIROLIMUS, PER 1 MG/ML  C9110 INJECTION, ALEMTUZUMAB, PER 10 MG/ ML  C9126 INJECTION, NATALIZUMAB, PER 5 MG  C9212 INJECTION, ALEFACEPT, FOR INTRAMUSCULAR USE, PER 7.5 MG  C9230 INJECTION, ABATACEPT, PER 10 MG  C9239 INJECTION, TEMSIROLIMUS, 1 MG  C9249 INJECTION, CERTOLIZUMAB PEGOL, 1 MG  C9261 INJECTION, USTEKINUMAB, 1 MG  C9264 INJECTION, TOCILIZUMAB, 1 MG  C9286 INJECTION, BELATACEPT, 1 MG  C9455 Injection, siltuximab, 10 mg  J0129 Injection, abatacept, 10 mg (code may be used for Medicare when drug administered under the direct supervision of a physician, not for use when drug is self  J0485 INJECTION, BELATACEPT, 1 MG  J0490 INJECTION, BELIMUMAB, 10 MG  J0718 INJECTION, CERTOLIZUMAB PEGOL, 1 MG  J1602 INJECTION, GOLIMUMAB, 1 MG, FOR INTRAVENOUS USE  J1745 INJECTION, INFLIXIMAB, EXCLUDES BIOSIMILAR, 10 MG  J2323 INJECTION, NATALIZUMAB, 1 MG  J2350 INJECTION, OCRELIZUMAB, 1 MG  J2860 INJECTION, SILTUXIMAB, 10 MG  J3262 INJECTION, TOCILIZUMAB, 1 MG  J7500 AZATHIOPRINE, ORAL, 50 MG  J7505 MUROMONAB  J7507 TACROLIMUS, IMMEDIATE RELEASE, ORAL, 1 MG  J7508 TACROLIMUS, EXTENDED RELEASE, (ASTAGRAF XL), ORAL, 0.1 MG  J7511 LYMPHOCYTE IMMUNE GLOBULIN, ANTITHYMOCYTE GLOBULIN, RABBIT, PARENTERAL, 25 MG  J7513 DACLIZUMAB, PARENTERAL, 25 MG  J7515 CYCLOSPORINE, ORAL, 25 MG  J7518 MYCOPHENOLIC ACID, ORAL, 180 MG  J7520 SIROLIMUS, ORAL, 1 MG  J7527 EVEROLIMUS, ORAL, 0.25 MG  J8610 Methotrexate, oral, 2.5 mg  J9010 ALEMTUZUMAB, 10 MG / INJECTION, ALEMTUZUMAB, 10 MG  J9091 Cyclophosphamide, 1. 0 gram / CYCLOPHOSPHAMIDE, 1.0 GRAM  J9092 Cyclophosphamide, 2. 0 gram / CYCLOPHOSPHAMIDE, 2.0 GRAM  J9094 CYCLOPHOSPHAMIDE, LYOPHILIZED, 200 MG  J9097 Cyclophosphamide, lyophilized, 2. 0 gram / CYCLOPHOSPHAMIDE, LYOPHILIZED, 2.0 GRAM  J9250 METHOTREXATE SODIUM, 5 MG  J9310 INJECTION, RITUXIMAB, 100 MG / RITUXIMAB, 100 MG  J9311 Injection, rituximab 10 mg and hyaluronidase  J9312 Injection, rituximab, 10 mg  J9330 INJECTION, TEMSIROLIMUS, 1 MG  K0412 MYCOPHENOLATE MOFETIL, ORAL, 250 MG  Q2019 INJECTION, BASILIXIMAB, 20 MG  Q4079 INJECTION, NATALIZUMAB, 1 MG  Q5103 INJECTION, INFLIXIMAB  S0087 ALEMTUZUMAB INJECTION, 30 MG  S0162 INJECTION, EFALIZUMAB, 125 MG  80180 Mycophenolate (mycophenolic acid)  C9211 INJECTION, ALEFACEPT, FOR INTRAVENOUS USE, PER 7.5 MG  C9219 MYCOPHENOLIC ACID, ORAL, PER 180 MG  C9236 INJECTION, ECULIZUMAB, 10 MG  C9419 INJECTION, CLADRIBINE, BRAND NAME, PER 1 MG  C9420 CYCLOPHOSPHAMIDE, BRAND NAME, 100 MG  C9421 CYCLOPHOSPHAMIDE, LYOPHILIZED, BRAND NAME, 100 MG  C9436 AZATHIOPRINE, PARENTERAL, BRAND NAME, PER 100 MG  C9438 CYCLOSPORINE, ORAL, BRAND NAME, 100 MG  J0135 INJECTION, ADALIMUMAB, 20 MG  J0202 INJECTION, ALEMTUZUMAB, 1 MG  J0215 INJECTION, ALEFACEPT, 0.5 MG  J0480 INJECTION, BASILIXIMAB, 20 MG  J0638 INJECTION, CANAKINUMAB, 1 MG  J0717 Injection, certolizumab pegol, 1 mg (code may be used for Medicare when drug administered under the direct supervision of a physician, not for use when drug is self  J1300 INJECTION, ECULIZUMAB, 10 MG  J1438 Injection, etanercept, 25 mg (code may be used for Medicare when drug administered under the direct supervision of a physician, not for use when drug is self  J1628 Injection, guselkumab, 1 mg  J2793 INJECTION, RILONACEPT, 1 MG  J3245 INJECTION, TILDRAKIZUMAB, 1 MG  J3357 USTEKINUMAB, FOR SUBCUTANEOUS INJECTION, 1 MG  J3358 USTEKINUMAB, FOR INTRAVENOUS INJECTION, 1 MG  J3380 INJECTION, VEDOLIZUMAB, 1 MG  J7501 AZATHIOPRINE, PARENTERAL, 100 MG  J7502 CYCLOSPORINE, ORAL, 100 MG  J7503 TACROLIMUS, EXTENDED RELEASE, (ENVARSUS XR), ORAL, 0.25 MG  J7504 LYMPHOCYTE IMMUNE GLOBULIN, ANTITHYMOCYTE GLOBULIN, EQUINE, PARENTERAL, 250 MG  J7517 MYCOPHENOLATE MOFETIL, ORAL, 250 MG  J7525 TACROLIMUS, PARENTERAL, 5 MG  J8530 Cyclophosphamide, oral, 25 mg  J8561 Everolimus, oral, 0. 25 mg / EVEROLIMUS, ORAL, 0.25 MG  J9065 INJECTION, CLADRIBINE, PER 1 MG  J9070 CYCLOPHOSPHAMIDE, 100 MG  J9080 CYCLOPHOSPHAMIDE, 200 MG  J9090 CYCLOPHOSPHAMIDE, 500 MG  J9093 CYCLOPHOSPHAMIDE, LYOPHILIZED, 100 MG  J9095 CYCLOPHOSPHAMIDE, LYOPHILIZED, 500 MG  J9096 Cyclophosphamide, lyophilized, 1. 0 gram / CYCLOPHOSPHAMIDE, LYOPHILIZED, 1.0 GRAM  J9260 METHOTREXATE SODIUM, 50 MG  K0119 AZATHIOPRINE  K0120 AZATHIOPRINE  K0121 CYCLOSPORINE  K0122 CYCLOSPORINE  K0123 LYMPHOCYTE IMMUNE GLOBULIN, ANTITHYMOCYTE GLOBULIN  Q2044 INJECTION, BELIMUMAB, 10 MG  Q5104 INJECTION, INFLIXIMAB  Q5109 INJECTION, INFLIXIMAB  S0193 INJECTION, ALEFACEPT, 7.5 MG (INCLUDES DOSE PACKAGING)  S9359 Home infusion therapy, antitumor necrosis factor intravenous therapy; (e.g., Infliximab); administrative services, professional pharmacy services, care coordination, and all necessary supplies and equipment (drugs and nursing visits coded separately), per diem |
| Cancer in the year prior to cancer therapy | [-365 prior to cancer therapy, start of cancer therapy] | 1 IP claim or 2 OP claims | Any | Cancer  ICD-10 diagnosis code  C00 Malignant neoplasm of lip  C02 Malignant neoplasm of other and unspecified parts of tongue  C03 Malignant neoplasm of gum  C09 Malignant neoplasm of tonsil  C10 Malignant neoplasm of oropharynx  C11 Malignant neoplasm of nasopharynx  C12 Malignant neoplasm of pyriform sinus  C15 Malignant neoplasm of esophagus  C17 Malignant neoplasm of small intestine  C18 Malignant neoplasm of colon  C20 Malignant neoplasm of rectum  C22 Malignant neoplasm of liver and intrahepatic bile ducts  C23 Malignant neoplasm of gallbladder  C25 Malignant neoplasm of pancreas  C31 Malignant neoplasm of accessory sinuses  C32 Malignant neoplasm of larynx  C38 Malignant neoplasm of heart, mediastinum and pleura  C39 Malignant neoplasm of other and ill  C41 Malignant neoplasm of bone and articular cartilage of other and unspecified sites  C43 Malignant melanoma of skin  C45 Mesothelioma  C46 Kaposi's sarcoma  C47 Malignant neoplasm of peripheral nerves and autonomic nervous system  C48 Malignant neoplasm of retroperitoneum and peritoneum  C50 Malignant neoplasm of breast  C51 Malignant neoplasm of vulva  C52 Malignant neoplasm of vagina  C53 Malignant neoplasm of cervix uteri  C54 Malignant neoplasm of corpus uteri  C55 Malignant neoplasm of uterus, part unspecified  C56 Malignant neoplasm of ovary  C57 Malignant neoplasm of other and unspecified female genital organs  C60 Malignant neoplasm of penis  C62 Malignant neoplasm of testis  C65 Malignant neoplasm of renal pelvis  C66 Malignant neoplasm of ureter  C68 Malignant neoplasm of other and unspecified urinary organs  C69 Malignant neoplasm of eye and adnexa  C73 Malignant neoplasm of thyroid gland  C74 Malignant neoplasm of adrenal gland  C76 Malignant neoplasm of other and ill  C82 Follicular lymphoma  C83 Non  C88 Malignant immunoproliferative diseases and certain other B  C90 Multiple myeloma and malignant plasma cell neoplasms  C91 Lymphoid leukemia  C92 Myeloid leukemia  C93 Monocytic leukemia  C95 Leukemia of unspecified cell type  C96 Other and unspecified malignant neoplasms of lymphoid, hematopoietic and related tissue  C01 Malignant neoplasm of base of tongue  C04 Malignant neoplasm of floor of mouth  C05 Malignant neoplasm of palate  C06 Malignant neoplasm of other and unspecified parts of mouth  C07 Malignant neoplasm of parotid gland  C08 Malignant neoplasm of other and unspecified major salivary glands  C13 Malignant neoplasm of hypopharynx  C14 Malignant neoplasm of other and ill  C16 Malignant neoplasm of stomach  C19 Malignant neoplasm of rectosigmoid junction  C21 Malignant neoplasm of anus and anal canal  C24 Malignant neoplasm of other and unspecified parts of biliary tract  C26 Malignant neoplasm of other and ill  C30 Malignant neoplasm of nasal cavity and middle ear  C33 Malignant neoplasm of trachea  C34 Malignant neoplasm of bronchus and lung  C37 Malignant neoplasm of thymus  C40 Malignant neoplasm of bone and articular cartilage of limbs  C49 Malignant neoplasm of other connective and soft tissue  C58 Malignant neoplasm of placenta  C61 Malignant neoplasm of prostate  C63 Malignant neoplasm of other and unspecified male genital organs  C64 Malignant neoplasm of kidney, except renal pelvis  C67 Malignant neoplasm of bladder  C70 Malignant neoplasm of meninges  C71 Malignant neoplasm of brain  C72 Malignant neoplasm of spinal cord, cranial nerves and other parts of central nervous system  C75 Malignant neoplasm of other endocrine glands and related structures  C81 Hodgkin lymphoma  C84 Mature T/NK  C85 Other specified and unspecified types of non  C94 Other leukemias of specified cell type |
| Cancer therapy | [-180, -1] |  |  |  |
| Any immunodeficiency | [start of data, -1] | 1 IP claim or 2 OP claims | Any | ICD-10 diagnosis code  D47.4 Osteomyelofibrosis  D47.9 Neoplasm of uncertain behavior of lymphoid, hematopoietic and related tissue, unspecified  D82 Immunodeficiency associated with other major defects  D82.0 Wiskott–Aldrich syndrome  D82.4 Hyperimmunoglobulin E [IgE] syndrome  D82.9 Immunodeficiency associated with major defect, unspecified  D45 Polycythemia vera  D46.22 Refractory anemia with excess of blasts 2  D47.1 Chronic myeloproliferative disease  D47.Z1 Post transplant lymphoproliferative disorder (PTLD)  D47.Z9 Other specified neoplasms of uncertain behavior of lymphoid, hematopoietic and related tissue  D61.82 Myelophthisis  D75.81 Myelofibrosis  D82.1 Di George's syndrome  D82.2 Immunodeficiency with short limbed stature  D82.3 Immunodeficiency following hereditary defective response to Epstein Barr virus  D82.8 Immunodeficiency associated with other specified major defects  R75 Inconclusive laboratory evidence of human immunodeficiency virus [HIV]  Z21 Asymptomatic human immunodeficiency virus [HIV] infection status |
| HIV infection | [start of data, -1] | 1 IP claim or 2 OP claims | Any | ICD-10 diagnosis code  B20 Human immunodeficiency virus [HIV] disease  B97.35 Human immunodeficiency virus, type 2 [HIV 2] as the cause of diseases classified elsewhere  R75 Inconclusive laboratory evidence of human immunodeficiency virus [HIV]  Z21 Asymptomatic human immunodeficiency virus [HIV] infection status  HCPCS/CPT  3490F History of AIDS defining condition (HIV) |
| Immunosuppressive therapy |  | 1 IP claim or 2 OP claims | Any | Generic name  BARICITINIB  BASILIXIMAB  BRODALUMAB  CANAKINUMAB/PF  DECITABINE/CEDAZURIDINE  ECULIZUMAB  FLUOROURACIL/ADHESIVE BANDAGE  GUSELKUMAB  INFLIXIMAB  INFLIXIMAB-ABDA  INFLIXIMAB-DYYB  NELARABINE  OCRELIZUMAB  PEMETREXED DISODIUM  PIRFENIDONE  PRALATREXATE  RAVULIZUMAB-CWVZ  ANAKINRA  BELATACEPT  CARMUSTINE IN POLIFEPROSAN 20  CERTOLIZUMAB PEGOL  CYCLOSPORINE/CHONDROITIN SULFATE A SODIUM  DACLIZUMAB  DAUNORUBICIN/CYTARABINE LIPOSOMAL  FINGOLIMOD HCL  GEMCITABINE HCL IN 0.9 % SODIUM CHLORIDE  GOLIMUMAB  INEBILIZUMAB-CDON  INFLIXIMAB-AXXQ  SATRALIZUMAB-MWGE  SECUKINUMAB  STREPTOZOCIN  TACROLIMUS IN VEHICLE BASE NO.238  TEPROTUMUMAB-TRBW  VEDOLIZUMAB  ABATACEPT  ABATACEPT/MALTOSE  ALEFACEPT  ALEMTUZUMAB  CYTARABINE LIPOSOME/PF  RISANKIZUMAB-RZAA  SARILUMAB  TACROLIMUS, MICRONIZED  TACROLIMUS/NIACINAMIDE  TERIFLUNOMIDE  USTEKINUMAB  AZATHIOPRINE SODIUM  DIROXIMEL FUMARATE  EFALIZUMAB  MELPHALAN HCL/BETADEX SULFOBUTYL ETHER SODIUM  MUROMONAB-CD3  MYCOPHENOLATE MOFETIL HCL  NATALIZUMAB  OZANIMOD HYDROCHLORIDE  RILONACEPT  TACROLIMUS/HYALURONATE SODIUM/NIACINAMIDE  TOCILIZUMAB  UPADACITINIB  APREMILAST  BELIMUMAB  BENDAMUSTINE HCL  FLOXURIDINE  SIPONIMOD  TEMSIROLIMUS  TILDRAKIZUMAB-ASMN  IXEKIZUMAB  POMALIDOMIDE  EMAPALUMAB-LZSG  OFATUMUMAB  TACROLIMUS ANHYDROUS  IFOSFAMIDE/MESNA  SILTUXIMAB  TOFACITINIB CITRATE  CLOFARABINE  LENALIDOMIDE  LOMUSTINE  CARMUSTINE  MELPHALAN  THALIDOMIDE  FLUDARABINE PHOSPHATE  CYTARABINE/PF  ETANERCEPT  AZACITIDINE  CLADRIBINE  THIOTEPA  CHLORAMBUCIL  ADALIMUMAB  IFOSFAMIDE  METHOTREXATE  MELPHALAN HCL  CYTARABINE  DECITABINE  DACARBAZINE  MYCOPHENOLATE SODIUM  DIMETHYL FUMARATE  MERCAPTOPURINE  BUSULFAN  CYCLOSPORINE, MODIFIED  LEFLUNOMIDE  SIROLIMUS  METHOTREXATE/PF  EVEROLIMUS  CAPECITABINE  CYCLOSPORINE  AZATHIOPRINE  GEMCITABINE HCL  METHOTREXATE SODIUM/PF  MYCOPHENOLATE MOFETIL  CYCLOPHOSPHAMIDE  METHOTREXATE SODIUM  FLUOROURACIL  TACROLIMUS  TEMOZOLOMIDE  HCPCS/CPT  80158 Cyclosporine  80169 Everolimus  80195 Sirolimus  80197 Tacrolimus  C9006 INJECTION, TACROLIMUS, PER 5 MG (1 AMP)  C9020 SIROLIMUS TABLET, 1 MG  C9026 Injection, vedolizumab, 1 mg  C9106 SIROLIMUS, PER 1 MG/ML  C9110 INJECTION, ALEMTUZUMAB, PER 10 MG/ ML  C9126 INJECTION, NATALIZUMAB, PER 5 MG  C9212 INJECTION, ALEFACEPT, FOR INTRAMUSCULAR USE, PER 7.5 MG  C9230 INJECTION, ABATACEPT, PER 10 MG  C9239 INJECTION, TEMSIROLIMUS, 1 MG  C9249 INJECTION, CERTOLIZUMAB PEGOL, 1 MG  C9261 INJECTION, USTEKINUMAB, 1 MG  C9264 INJECTION, TOCILIZUMAB, 1 MG  C9286 INJECTION, BELATACEPT, 1 MG  C9455 Injection, siltuximab, 10 mg  J0129 Injection, abatacept, 10 mg (code may be used for Medicare when drug administered under the direct supervision of a physician, not for use when drug is self  J0485 INJECTION, BELATACEPT, 1 MG  J0490 INJECTION, BELIMUMAB, 10 MG  J0718 INJECTION, CERTOLIZUMAB PEGOL, 1 MG  J1602 INJECTION, GOLIMUMAB, 1 MG, FOR INTRAVENOUS USE  J1745 INJECTION, INFLIXIMAB, EXCLUDES BIOSIMILAR, 10 MG  J2323 INJECTION, NATALIZUMAB, 1 MG  J2350 INJECTION, OCRELIZUMAB, 1 MG  J2860 INJECTION, SILTUXIMAB, 10 MG  J3262 INJECTION, TOCILIZUMAB, 1 MG  J7500 AZATHIOPRINE, ORAL, 50 MG  J7505 MUROMONAB  J7507 TACROLIMUS, IMMEDIATE RELEASE, ORAL, 1 MG  J7508 TACROLIMUS, EXTENDED RELEASE, (ASTAGRAF XL), ORAL, 0.1 MG  J7511 LYMPHOCYTE IMMUNE GLOBULIN, ANTITHYMOCYTE GLOBULIN, RABBIT, PARENTERAL, 25 MG  J7513 DACLIZUMAB, PARENTERAL, 25 MG  J7515 CYCLOSPORINE, ORAL, 25 MG  J7518 MYCOPHENOLIC ACID, ORAL, 180 MG  J7520 SIROLIMUS, ORAL, 1 MG  J7527 EVEROLIMUS, ORAL, 0.25 MG  J8610 Methotrexate, oral, 2.5 mg  J9010 ALEMTUZUMAB, 10 MG / INJECTION, ALEMTUZUMAB, 10 MG  J9091 Cyclophosphamide, 1. 0 gram / CYCLOPHOSPHAMIDE, 1.0 GRAM  J9092 Cyclophosphamide, 2. 0 gram / CYCLOPHOSPHAMIDE, 2.0 GRAM  J9094 CYCLOPHOSPHAMIDE, LYOPHILIZED, 200 MG  J9097 Cyclophosphamide, lyophilized, 2. 0 gram / CYCLOPHOSPHAMIDE, LYOPHILIZED, 2.0 GRAM  J9250 METHOTREXATE SODIUM, 5 MG  J9310 INJECTION, RITUXIMAB, 100 MG / RITUXIMAB, 100 MG  J9311 Injection, rituximab 10 mg and hyaluronidase  J9312 Injection, rituximab, 10 mg  J9330 INJECTION, TEMSIROLIMUS, 1 MG  K0412 MYCOPHENOLATE MOFETIL, ORAL, 250 MG  Q2019 INJECTION, BASILIXIMAB, 20 MG  Q4079 INJECTION, NATALIZUMAB, 1 MG  Q5103 INJECTION, INFLIXIMAB  S0087 ALEMTUZUMAB INJECTION, 30 MG  S0162 INJECTION, EFALIZUMAB, 125 MG  80180 Mycophenolate (mycophenolic acid)  C9211 INJECTION, ALEFACEPT, FOR INTRAVENOUS USE, PER 7.5 MG  C9219 MYCOPHENOLIC ACID, ORAL, PER 180 MG  C9236 INJECTION, ECULIZUMAB, 10 MG  C9419 INJECTION, CLADRIBINE, BRAND NAME, PER 1 MG  C9420 CYCLOPHOSPHAMIDE, BRAND NAME, 100 MG  C9421 CYCLOPHOSPHAMIDE, LYOPHILIZED, BRAND NAME, 100 MG  C9436 AZATHIOPRINE, PARENTERAL, BRAND NAME, PER 100 MG  C9438 CYCLOSPORINE, ORAL, BRAND NAME, 100 MG  J0135 INJECTION, ADALIMUMAB, 20 MG  J0202 INJECTION, ALEMTUZUMAB, 1 MG  J0215 INJECTION, ALEFACEPT, 0.5 MG  J0480 INJECTION, BASILIXIMAB, 20 MG  J0638 INJECTION, CANAKINUMAB, 1 MG  J0717 Injection, certolizumab pegol, 1 mg (code may be used for Medicare when drug administered under the direct supervision of a physician, not for use when drug is self  J1300 INJECTION, ECULIZUMAB, 10 MG  J1438 Injection, etanercept, 25 mg (code may be used for Medicare when drug administered under the direct supervision of a physician, not for use when drug is self  J1628 Injection, guselkumab, 1 mg  J2793 INJECTION, RILONACEPT, 1 MG  J3245 INJECTION, TILDRAKIZUMAB, 1 MG  J3357 USTEKINUMAB, FOR SUBCUTANEOUS INJECTION, 1 MG  J3358 USTEKINUMAB, FOR INTRAVENOUS INJECTION, 1 MG  J3380 INJECTION, VEDOLIZUMAB, 1 MG  J7501 AZATHIOPRINE, PARENTERAL, 100 MG  J7502 CYCLOSPORINE, ORAL, 100 MG  J7503 TACROLIMUS, EXTENDED RELEASE, (ENVARSUS XR), ORAL, 0.25 MG  J7504 LYMPHOCYTE IMMUNE GLOBULIN, ANTITHYMOCYTE GLOBULIN, EQUINE, PARENTERAL, 250 MG  J7517 MYCOPHENOLATE MOFETIL, ORAL, 250 MG  J7525 TACROLIMUS, PARENTERAL, 5 MG  J8530 Cyclophosphamide, oral, 25 mg  J8561 Everolimus, oral, 0. 25 mg / EVEROLIMUS, ORAL, 0.25 MG  J9065 INJECTION, CLADRIBINE, PER 1 MG  J9070 CYCLOPHOSPHAMIDE, 100 MG  J9080 CYCLOPHOSPHAMIDE, 200 MG  J9090 CYCLOPHOSPHAMIDE, 500 MG  J9093 CYCLOPHOSPHAMIDE, LYOPHILIZED, 100 MG  J9095 CYCLOPHOSPHAMIDE, LYOPHILIZED, 500 MG  J9096 Cyclophosphamide, lyophilized, 1. 0 gram / CYCLOPHOSPHAMIDE, LYOPHILIZED, 1.0 GRAM  J9260 METHOTREXATE SODIUM, 50 MG  K0119 AZATHIOPRINE  K0120 AZATHIOPRINE  K0121 CYCLOSPORINE  K0122 CYCLOSPORINE  K0123 LYMPHOCYTE IMMUNE GLOBULIN, ANTITHYMOCYTE GLOBULIN  Q2044 INJECTION, BELIMUMAB, 10 MG  Q5104 INJECTION, INFLIXIMAB  Q5109 INJECTION, INFLIXIMAB  S0193 INJECTION, ALEFACEPT, 7.5 MG (INCLUDES DOSE PACKAGING)  S9359 Home infusion therapy, antitumor necrosis factor intravenous therapy; (e.g., Infliximab); administrative services, professional pharmacy services, care coordination, and all necessary supplies and equipment (drugs and nursing visits coded separately), per diem |

CPT, current procedural terminology; ICD-10, International Statistical Classification of Diseases and Related Health Problems, 10^th^ revision.

**Table S4. Pre- and post-weighting characteristics for the 3-dose homologous series subgroup.**

|  | **Pre-PS weighting baseline characteristics** | | | **Post-PS weighting (truncated at 99th percentile)** | | |
| --- | --- | --- | --- | --- | --- | --- |
|  | **Third dose of mRNA-1273** | **Third dose of BNT162b2** | **Weighted absolute standardized difference** | **Third dose of mRNA-1273** | **Third dose of BNT162b2** | **Weighted absolute standardized difference** |
| **Number of patients** | 90,984 | 85,619 |  | 90,984 | 85,619 |  |
| Sum of weights |  |  |  | 170,832.71 | 175,804.86 |  |
| Effective sample size |  |  |  | 80,513 | 73,864 |  |
| **Age (continuous)** |  |  | 0.013 |  |  | 0.000 |
| ...Mean (SD), years | 71.65 (7.76) | 71.55 (8.04) |  | 71.44 (7.74) | 71.44 (7.81) |  |
| ...Median [IQR], years | 69.00 [66.00, 74.00] | 69.00  [66.00, 74.00] |  |  |  |  |
| **Age (categorical)** |  |  |  |  |  |  |
| ...65-74 years | 69,235 (76.1%) | 65,470 (76.5%) | 0.009 | 131,522.6 (77.0%) | 135,503.1 (77.1%) | 0.002 |
| ...85-84 years | 14,976 (16.5%) | 13,182 (15.4%) | 0.029 | 26,759.2 (15.7%) | 27,246.6 (15.5%) | 0.005 |
| ...85+ years | 6773 (7.4%) | 6,967 (8.1%) | 0.026 | 12,550.9 (7.4%) | 13,055.1 (7.4%) | 0.003 |
| **Sex** |  |  |  |  |  |  |
| ...Female | 48,758 (53.6%) | 45,958 (53.7%) | 0.002 | 91,609.4 (53.6%) | 94,087.5 (53.5%) | 0.002 |
| ...Male | 42,226 (46.4%) | 39,661 (46.3%) | 0.002 | 79,223.3 (46.4%) | 81,717.4 (46.5%) | 0.002 |
| **Primary payer type** |  |  |  |  |  |  |
| ...Missing | 3097 (3.4%) | 4178 (4.9%) | 0.074 | 6482.8 (3.8%) | 7052.1 (4.0%) | 0.011 |
| ...Commercial | 38,646 (42.5%) | 42,439 (49.6%) | 0.143 | 79,074.5 (46.3%) | 82,255.7 (46.8%) | 0.010 |
| ...Medicare | 27,007 (29.7%) | 21,276 (24.8%) | 0.109 | 46,447.7 (27.2%) | 47,309.7 (26.9%) | 0.006 |
| ...Medicaid | 22,234 (24.4%) | 17,726 (20.7%) | 0.089 | 38,827.6 (22.7%) | 39,187.3 (22.3%) | 0.011 |
| **State** |  |  |  |  |  |  |
| ...Alaska/Other/ Puerto Rico | 130 (0.1%) | 40 (0.0%) | 0.031 | 166.0 (0.1%) | 145.4 (0.1%) | 0.005 |
| ...Alabama | 87 (0.1%) | 71 (0.1%) | 0.004 | 157.5 (0.1%) | 157.7 (0.1%) | 0.001 |
| ...Arkansas | 2322 (2.6%) | 2324 (2.7%) | 0.010 | 4387.9 (2.6%) | 4618.8 (2.6%) | 0.004 |
| ...Arizona | 12,921 (14.2%) | 4985 (5.8%) | 0.282 | 17,734.0 (10.4%) | 17,068.4 (9.7%) | 0.022 |
| ...California | 10,170 (11.2%) | 5246 (6.1%) | 0.180 | 15,255.0 (8.9%) | 15,362.3 (8.7%) | 0.007 |
| ...Colorado | 264 (0.3%) | 298 (0.3%) | 0.010 | 540.7 (0.3%) | 559.8 (0.3%) | 0.000 |
| ...Connecticut | 465 (0.5%) | 817 (1.0%) | 0.052 | 1255.1 (0.7%) | 1287.9 (0.7%) | 0.000 |
| ...DC | 177 (0.2%) | 92 (0.1%) | 0.022 | 267.8 (0.2%) | 264.3 (0.2%) | 0.002 |
| ...Delaware | 264 (0.3%) | 442 (0.5%) | 0.036 | 648.0 (0.4%) | 702.5 (0.4%) | 0.003 |
| ...Florida | 2031 (2.2%) | 2,005 (2.3%) | 0.007 | 3944.6 (2.3%) | 4051.9 (2.3%) | 0.000 |
| ...Georgia | 1015 (1.1%) | 592 (0.7%) | 0.045 | 1634.6 (1.0%) | 1654.7 (0.9%) | 0.002 |
| ...Hawaii | 130 (0.1%) | 105 (0.1%) | 0.006 | 237.7 (0.1%) | 241.5 (0.1%) | 0.001 |
| ...Iowa | 702 (0.8%) | 352 (0.4%) | 0.047 | 1063.6 (0.6%) | 1102.4 (0.6%) | 0.001 |
| ...Idaho/ Wyoming | 1300 (1.4%) | 1,809 (2.1%) | 0.052 | 2813.0 (1.7%) | 3126.8 (1.8%) | 0.010 |
| ...Illinois | 9053 (10.0%) | 11,406 (13.3%) | 0.105 | 20,282.3 (11.9%) | 20,679.1 (11.8%) | 0.003 |
| ...Indiana | 462 (0.5%) | 777 (0.9%) | 0.048 | 1172.1 (0.7%) | 1239.5 (0.7%) | 0.002 |
| ...Kansas | 1,356 (1.5%) | 2,308 (2.7%) | 0.084 | 3333.1 (2.0%) | 3634.9 (2.1%) | 0.008 |
| ...Kentucky | 265 (0.3%) | 440 (0.5%) | 0.035 | 695.2 (0.4%) | 717.0 (0.4%) | 0.000 |
| ...Louisiana | 450 (0.5%) | 351 (0.4%) | 0.013 | 795.1 (0.5%) | 807.0 (0.5%) | 0.001 |
| ...Massachusetts/Rhode Island | 312 (0.3%) | 323 (0.4%) | 0.006 | 625.8 (0.4%) | 639.1 (0.4%) | 0.001 |
| ...Maryland | 262 (0.3%) | 226 (0.3%) | 0.005 | 477.3 (0.3%) | 486.1 (0.3%) | 0.001 |
| ...Maine | 50 (0.1%) | 73 (0.1%) | 0.011 | 116.6 (0.1%) | 122.1 (0.1%) | 0.001 |
| ...Michigan | 9331 (10.3%) | 10,513 (12.3%) | 0.064 | 18,617.9 (10.9%) | 19,762.8 (11.2%) | 0.011 |
| ...Minnesota | 140 (0.2%) | 150 (0.2%) | 0.005 | 289.9 (0.2%) | 296.6 (0.2%) | 0.000 |
| ...Missouri | 243 (0.3%) | 308 (0.4%) | 0.017 | 545.6 (0.3%) | 553.3 (0.3%) | 0.001 |
| ...Mississippi | 101 (0.1%) | 59 (0.1%) | 0.014 | 165.5 (0.1%) | 167.0 (0.1%) | 0.001 |
| ...Montana | 195 (0.2%) | 191 (0.2%) | 0.002 | 387.2 (0.2%) | 390.6 (0.2%) | 0.001 |
| ...North/South Dakota | 32 (0.0%) | 54 (0.1%) | 0.013 | 76.8 (0.0%) | 83.8 (0.1%) | 0.001 |
| ...North Carolina | 319 (0.4%) | 449 (0.5%) | 0.026 | 727.6 (0.4%) | 771.2 (0.4%) | 0.002 |
| ...Nebraska | 282 (0.3%) | 622 (0.7%) | 0.058 | 822.4 (0.5%) | 899.0 (0.5%) | 0.004 |
| ...New Hampshire | 30 (0.0%) | 34 (0.0%) | 0.004 | 68.3 (0.0%) | 65.3 (0.0%) | 0.001 |
| ...New Jersey | 5014 (5.5%) | 7,533 (8.8%) | 0.128 | 11,986.4 (7.0%) | 12,557.2 (7.1%) | 0.005 |
| ...New Mexico | 1307 (1.4%) | 955 (1.1%) | 0.029 | 2195.6 (1.3%) | 2234.4 (1.3%) | 0.001 |
| ...Nevada | 205 (0.2%) | 238 (0.3%) | 0.011 | 425.3 (0.3%) | 439.7 (0.3%) | 0.000 |
| ...New York | 3155 (3.5%) | 1603 (1.9%) | 0.099 | 4734.4 (2.8%) | 4683.6 (2.7%) | 0.007 |
| ...Ohio | 5927 (6.5%) | 6723 (7.9%) | 0.052 | 11,948.9 (7.0%) | 12,523.9 (7.1%) | 0.005 |
| ...Oklahoma | 694 (0.8%) | 927 (1.1%) | 0.033 | 1510.8 (0.9%) | 1622.2 (0.9%) | 0.004 |
| ...Oregon | 338 (0.4%) | 215 (0.3%) | 0.022 | 540.8 (0.3%) | 528.4 (0.3%) | 0.003 |
| ...Pennsylvania | 3315 (3.6%) | 2571 (3.0%) | 0.036 | 5930.4 (3.5%) | 5986.1 (3.4%) | 0.004 |
| ...South Carolina | 186 (0.2%) | 171 (0.2%) | 0.001 | 335.4 (0.2%) | 347.8 (0.2%) | 0.000 |
| ...Tennessee | 180 (0.2%) | 162 (0.2%) | 0.002 | 349.8 (0.2%) | 357.0 (0.2%) | 0.000 |
| ...Texas | 5443 (6.0%) | 6593 (7.7%) | 0.068 | 11,967.0 (7.0%) | 12,313.2 (7.0%) | 0.000 |
| ...Utah | 36 (0.0%) | 37 (0.0%) | 0.002 | 73.9 (0.0%) | 75.7 (0.0%) | 0.000 |
| ...Virginia | 3491 (3.8%) | 3018 (3.5%) | 0.017 | 6460.8 (3.8%) | 6621.7 (3.8%) | 0.001 |
| ...Vermont | 79 (0.1%) | 80 (0.1%) | 0.002 | 152.0 (0.1%) | 157.4 (0.1%) | 0.000 |
| ...Washington | 899 (1.0%) | 664 (0.8%) | 0.023 | 1523.5 (0.9%) | 1563.9 (0.9%) | 0.000 |
| ...Wisconsin | 5816 (6.4%) | 6637 (7.8%) | 0.053 | 11,328.6 (6.6%) | 12,067.9 (6.9%) | 0.009 |
| ...West Virginia | 38 (0.0%) | 30 (0.0%) | 0.003 | 65.0 (0.0%) | 66.0 (0.0%) | 0.000 |
| **Month and year of index** |  |  |  |  |  |  |
| ...September 2021 | 619 (0.7%) | 8127 (9.5%) | 0.409 | 3137.6 (1.8%) | 8743.8 (5.0%) | 0.174 |
| ...October 2021 | 14,806 (16.3%) | 28,816 (33.7%) | 0.41 | 43,448.1 (25.4%) | 43,758.5 (24.9%) | 0.013 |
| ...November 2021 | 32,037 (35.2%) | 16,300 (19.0%) | 0.37 | 48,291.1 (28.3%) | 47,621.3 (27.1%) | 0.026 |
| ...December 2021 | 17,982 (19.8%) | 12,300 (14.4%) | 0.144 | 30,317.4 (17.8%) | 30,317.9 (17.3%) | 0.013 |
| ...January 2022 | 9042 (9.9%) | 6798 (7.9%) | 0.07 | 15,843.1 (9.3%) | 15,790.3 (9.0%) | 0.010 |
| ...February 2022 | 2581 (2.8%) | 2160 (2.5%) | 0.019 | 4755.1 (2.8%) | 4737.5 (2.7%) | 0.005 |
| ...March 2022 | 1577 (1.7%) | 1288 (1.5%) | 0.018 | 2877.6 (1.7%) | 2860.4 (1.6%) | 0.005 |
| ...April 2022 | 4923 (5.4%) | 3598 (4.2%) | 0.057 | 8502.2 (5.0%) | 8411.0 (4.8%) | 0.009 |
| ...May 2022 | 3082 (3.4%) | 2453 (2.9%) | 0.03 | 5516.1 (3.2%) | 5464.5 (3.1%) | 0.007 |
| ...June 2022 | 1744 (1.9%) | 1429 (1.7%) | 0.019 | 3174.4 (1.9%) | 3151.6 (1.8%) | 0.005 |
| ...July 2022 | 1967 (2.2%) | 1781 (2.1%) | 0.006 | 3755.6 (2.2%) | 3736.9 (2.1%) | 0.005 |
| ...August 2022 | 624 (0.7%) | 569 (0.7%) | 0.003 | 1214.5 (0.7%) | 1211.0 (0.7%) | 0.003 |
| **Count of unique NDC generic names** |  |  | 0.065 |  |  | 0.009 |
| ...Mean (SD) | 8.15 (6.43) | 7.73 (6.30) |  | 7.99 (6.30) | 7.93 (6.46) |  |
| ...Median [IQR] | 7.00 [4.00, 11.00] | 6.00 [3.00, 11.00] |  | 7.00 [4.00, 11.00] | 6.00 [3.00, 11.00] |  |
| **Count of hospitalization events in 365-day baseline period** |  |  | 0.032 |  |  | 0.001 |
| ...Mean (SD) | 0.61 (3.74) | 0.74 (4.34) |  | 0.69 (4.57) | 0.69 (3.87) |  |
| ...Median [IQR] | 0.00 [0.00, 0.00] | 0.00 [0.00, 0.00] |  | 0.00 [0.00, 0.00] | 0.00 [0.00, 0.00] |  |
| **Count of outpatient events in 365-day baseline period** |  |  | 0.026 |  |  | 0.000 |
| ...Mean (SD) | 35.86 (69.92) | 37.69 (71.99) |  | 36.31 (70.80) | 36.28 (69.90) |  |
| ...Median [IQR] | 15.00 [8.00, 28.00] | 15.00 [8.00, 30.00] |  | 15.00  [8.00, 28.00] | 15.00  [8.00, 30.00] |  |
| **Comorbid conditions and scores** |  |  |  |  |  |  |
| **Charlson comorbidity score categories** |  |  |  |  |  |  |
| ...0 | 37,893 (41.6%) | 36,924 (43.1%) | 0.030 | 72,620.8 (42.5%) | 75,016.7 (42.7%) | 0.003 |
| ...1 | 17,485 (19.2%) | 16,362 (19.1%) | 0.003 | 32,621.7 (19.1%) | 33,604.6 (19.1%) | 0.001 |
| ...2+ | 35,606 (39.1%) | 32,333 (37.8%) | 0.028 | 65,590.2 (38.4%) | 67,183.5 (38.2%) | 0.004 |
| **Frailty score categories** |  |  |  |  |  |  |
| ...Robust (0-0.149) | 63,569 (69.9%) | 59,974 (70.0%) | 0.004 | 119,960.9 (70.2%) | 123,548.0 (70.3%) | 0.001 |
| ...Prefrail (0.15-0.249) | 24,136 (26.5%) | 21,840 (25.5%) | 0.023 | 44,333.3 (26.0%) | 45,425.1 (25.8%) | 0.003 |
| ...Mild frailty (0.25-0.349) | 2841 (3.1%) | 3,273 (3.8%) | 0.038 | 5621.1 (3.3%) | 5883.9 (3.4%) | 0.003 |
| ...Moderate to severe frailty (≥0.35) | 438 (0.5%) | 532 (0.6%) | 0.019 | 917.4 (0.5%) | 947.9 (0.5%) | 0.000 |
| **Clinical conditions** |  |  |  |  |  |  |
| Alcohol use; n (%) | 2767 (3.0%) | 2433 (2.8%) | 0.012 | 5089.3 (3.0%) | 5223.8 (3.0%) | 0.001 |
| Arrhythmia; n (%) | 22,459 (24.7%) | 21,375 (25.0%) | 0.006 | 42,129.8 (24.7%) | 43,351.6 (24.7%) | 0.000 |
| Asthma; n (%) | 9,196 (10.1%) | 8,747 (10.2%) | 0.004 | 17,383.0 (10.2%) | 17,958.7 (10.2%) | 0.001 |
| Cancer; n (%) | 11,188 (12.3%) | 10,897 (12.7%) | 0.013 | 21,426.4 (12.5%) | 22,011.9 (12.5%) | 0.001 |
| Cardiovascular disease; n (%) | 71,625 (78.7%) | 66,327 (77.5%) | 0.030 | 133,172.6 (78.0%) | 136,884.2 (77.9%) | 0.002 |
| Cerebrovascular disease; n (%) | 11,318 (12.4%) | 10,709 (12.5%) | 0.002 | 21,215.6 (12.4%) | 21,792.4 (12.4%) | 0.001 |
| Coronary artery disease; n (%) | 17,449 (19.2%) | 16,507 (19.3%) | 0.003 | 32,686.2 (19.1%) | 33,641.8 (19.1%) | 0.000 |
| Chronic kidney disease; n (%) | 16,210 (17.8%) | 14,386 (16.8%) | 0.027 | 29,412.7 (17.2%) | 30,107.4 (17.1%) | 0.002 |
| Chronic lung disease; n (%) | 18,850 (20.7%) | 17,209 (20.1%) | 0.015 | 35,007.2 (20.5%) | 35,973.8 (20.5%) | 0.001 |
| COPD; n (%) | 12,427 (13.7%) | 10,927 (12.8%) | 0.026 | 22,736.7 (13.3%) | 23,263.6 (13.2%) | 0.002 |
| Dementia; n (%) | 3706 (4.1%) | 4474 (5.2%) | 0.055 | 7187.9 (4.2%) | 7687.6 (4.4%) | 0.008 |
| Diabetes; n (%) | 28,491 (31.3%) | 25,222 (29.5%) | 0.040 | 52,028.6 (30.5%) | 53,171.1 (30.2%) | 0.005 |
| Down syndrome¹; n (%) | 17 (0.0%) | 15 (0.0%) | 0.001 | 35.3 (0.0%) | 26.5 (0.0%) | 0.004 |
| Congestive heart failure; n (%) | 9,269 (10.2%) | 8,890 (10.4%) | 0.006 | 17,401.4 (10.2%) | 17,844.9 (10.2%) | 0.001 |
| Hypertension; n (%) | 64,567 (71.0%) | 59,501 (69.5%) | 0.032 | 119,719.3 (70.1%) | 122,969.1 (70.0%) | 0.003 |
| Irritable bowel syndrome; n (%) | 27,309 (30.0%) | 25,465 (29.7%) | 0.006 | 50,843.5 (29.8%) | 52,281.2 (29.7%) | 0.001 |
| Liver disease; n (%) | 6823 (7.5%) | 5694 (6.7%) | 0.033 | 12,293.0 (7.2%) | 12,514.3 (7.1%) | 0.003 |
| Obesity; n (%) | 24,843 (27.3%) | 22,110 (25.8%) | 0.034 | 45,587.0 (26.7%) | 46,710.3 (26.6%) | 0.003 |
| Influenza or RSV; n (%) | 513 (0.6%) | 566 (0.7%) | 0.012 | 1026.1 (0.6%) | 1070.0 (0.6%) | 0.001 |
| Psoriasis; n (%) | 806 (0.9%) | 781 (0.9%) | 0.003 | 1558.0 (0.9%) | 1600.1 (0.9%) | 0.000 |
| Psoriatic arthritis; n (%) | 355 (0.4%) | 341 (0.4%) | 0.001 | 703.3 (0.4%) | 723.6 (0.4%) | 0.000 |
| Rheumatoid arthritis; n (%) | 2149 (2.4%) | 1,835 (2.1%) | 0.015 | 3931.7 (2.3%) | 3991.2 (2.3%) | 0.002 |
| Sickle cell disease or thalassemia; n (%) | 78 (0.1%) | 67 (0.1%) | 0.003 | 136.5 (0.1%) | 139.9 (0.1%) | 0.000 |
| History of tobacco use/smoking; n (%) | 22,121 (24.3%) | 20,299 (23.7%) | 0.014 | 41,092.0 (24.1%) | 42,258.5 (24.0%) | 0.000 |

COPD, chronic obstructive pulmonary disease; IQR, interquartile range; NDC, national drug code; PS, propensity score; SD, standard deviation.

**Table S5. COVID-19 hospitalization sensitivity analysis effect estimate.**

| **No. of events** | | **Weighted no. of events** | | **Weighted rate (95% CI) (per 1000 PYs)** | | **HR (95% CI)** | | **Weighted median follow-up time; days [IQR]** | | **Weighted time to event; median days to event among those with an event [IQR]** | |
| --- | --- | --- | --- | --- | --- | --- | --- | --- | --- | --- | --- |
| **Third dose of mRNA-1273** | **Third dose of BNT162b2** | **Third dose of mRNA-1273** | **Third dose of BNT162b2** | **Third dose of mRNA-1273** | **Third dose of BNT162b2** | **HR (95% CI)** | **P value** | **Third dose of mRNA-1273** | **Third dose of BNT162b2** | **Third dose of mRNA-1273** | **Third dose of BNT162b2** |
| **Primary analysis: COVID-19 hospitalization** | | | | | | | | | | | |
| 262 | 378 | 517.468 | 726.945 | 5.61 (5.13, 6.09) | 7.06 (6.54, 7.57) | 0.82 (0.69, 0.98) | 0.029 | 195.70 [136.52, 252.93] | 220.86 [144.62, 269.83] | 112.52 [58.48, 183.48] | 147.88 [65.49, 214.18] |
| **Sensitivity analysis: COVID-19 hospitalization identified in any diagnosis position** | | | | | | | | | | | |
| 382 | 538 | 739.107 | 1028.596 | 8.02 (7.44, 8.60) | 9.99 (9.38, 10.60) | 0.83 (0.72, 0.96) | 0.013 | 195.58 [136.37, 252.85] | 220.60 [144.25, 269.68] | 105.79 [51.97, 193.87] | 146.42 [70.83, 206.85] |
| **Sensitivity analysis: COVID-19 hospitalization captured with a more restrictive respiratory distress definition** | | | | | | | | | | | |
| 208 | 305 | 419.662 | 585.406 | 4.55 (4.11, 4.98) | 5.68 (5.22, 6.14) | 0.83 (0.68, 1.02) | 0.072 | 195.76 [136.59, 252.96] | 220.98 [144.77, 269.89] | 104.66 [53.15, 181.77] | 137.74 [65.14, 209.65] |
| **Sensitivity analysis: COVID-19 hospitalization captured using less restrictive cohort entry requirement*** | | | | | | | | | | | |
| 269 | 391 | 528.172 | 753.601 | 5.61 (5.14, 6.09) | 7.18 (6.67, 7.70) | 0.81 (0.68, 0.96) | 0.015 | 192.94 [133.82, 252.12] | 218.26 [138.77, 268.55] | 111.8 [57.41, 183.49] | 139.64 [65.16, 211.36] |
| **Sensitivity analysis: COVID-19 hospitalization captured within a cohort constructed using either open or closed claims where all patients have evidence of closed claims enrollment** | | | | | | | | | | | |
| 880 | 1,036 | 1,586.185 | 2,120.468 | 10.90 (10.36, 11.44) | 13.47 (12.90, 14.05) | 0.84 (0.76, 0.93) | 0.001 | 176.98 [118.59, 246.07] | 200.85 [119.10, 258.50] | 116.21 [57.33, 196.39] | 128.04 [67.93, 204.37] |
| **Sensitivity analysis: COVID-19 hospitalization captured within a cohort constructed using a 180-day COVID-19 washout period** | | | | | | | | | | | |
| 290 | 454 | 581.877 | 852.231 | 5.82 (5.35, 6.29) | 7.64 (7.13, 8.16) | 0.79 (0.67, 0.93) | 0.004 | 195.92 [136.06, 252.80] | 220.33 [143.10, 269.33] | 105.72 [59.12, 181.49] | 124.22 [64.4, 207.33] |

CI, confidence interval; HR, hazard ratio; IQR, interquartile range; PY, person-year.

*Less restrictive cohort entry requirement does not exclude patients who had an additional vaccine record within 42 days of their second dose or before September 22, 2021. Patients in the cohort are still required to have their qualifying event occur after September 22, 2021, and after 42 days following their second dose

**Table S6. Medically attended COVID-19 sensitivity analysis effect estimate.**

| **No. of events** | | **Weighted no. of events** | | **Weighted rate (95% CI) (per 1000 py)** | | **HR (95% CI)** | | **Weighted median follow-up time; days [IQR]** | | **Weighted time to event; median days to event among those with an event [IQR]** | |
| --- | --- | --- | --- | --- | --- | --- | --- | --- | --- | --- | --- |
| **Third dose of mRNA-1273** | **Third dose of BNT162b2** | **Third dose of mRNA-1273** | **Third dose of BNT162b2** | **Third dose of mRNA-1273** | **Third dose of BNT162b2** | **HR (95% CI)** | **P value** | **Third dose of mRNA-1273** | **Third dose of BNT162b2** | **Third dose of mRNA-1273** | **Third dose of BNT162b2** |
| **Primary analysis: medically attended COVID-19** | | | | | | | | | | | |
| 4401 | 5505 | 8564.205 | 10667.882 | 95.05 (93.03, 97.06) | 106.55 (104.53, 108.57) | 0.93 (0.89, 0.98) | 0.003 | 189.60 [132.12, 250.07] | 214.70 [135.18, 265.84] | 137.18 [59.29, 201.9] | 140.21 [63.12, 207.34] |
| **Sensitivity analysis: medically attended COVID-19 captured using less restrictive cohort entry requirement*** | | | | | | | | | | | |
| 4590 | 5673 | 8,924.874 | 11,014.532 | 97.16 (95.14, 99.18) | 108.04 (106.03, 110.06) | 0.94 (0.90, 0.98) | 0.005 | 187.02 [129.57, 249.17] | 211.78 [132.03, 264.16] | 131.25 [57.94, 199.98] | 135.77 [62.5, 206] |
| **Sensitivity analysis: medically attended COVID-19 captured within a cohort constructed using either open or closed claims where all patients have evidence of closed claims enrollment** | | | | | | | | | | | |
| 7137 | 7459 | 12,706.224 | 15,940.251 | 89.01 (87.46, 90.56) | 103.79 (102.18, 105.40) | 0.89 (0.86, 0.92) | 0.000 | 172.93 [113.95, 243.23] | 194.54 [113.21, 254.95] | 128.39 [57.56, 198.98] | 124.23 [60.09, 200.54] |
| **Sensitivity analysis: medically attended COVID-19 captured within a cohort constructed using a 180-day COVID-19 washout period** | | | | | | | | | | | |
| 4863 | 6235 | 9535.672 | 11,963.273 | 97.75 (95.79, 99.71) | 110.57 (108.58, 112.55) | 0.92 (0.89, 0.96) | 0.000 | 189.58 [131.37, 249.93] | 214.20 [133.90, 265.26] | 133.84 [58.27, 200.22] | 135.38 [62.08, 206.25] |

CI, confidence interval; HR, hazard ratio; IQR, interquartile range; PY, person-year.

*Less restrictive cohort entry requirement does not exclude patients who had an additional vaccine record within 42 days of their second dose or before September 22, 2021. Patients in the cohort are still required to have their qualifying event occur after September 22, 2021, and after 42 days following their second dose

Supplementary details

Supplementary file 1. Timing between first and second dose in a primary series and primary series to third dose

Supplementary file 2. Details on defining COVID-19 hospitalization

Supplementary file 3. Defining respiratory distress

**Supplementary file 1. Timing between first and second dose in a primary series and primary series to third dose**

**Minimum and maximum interval between a first dose and a second dose in a primary series**

An exploratory analysis was completed to assess the minimal interval that should be applied to define a second dose following an observed first dose for a heterologous vaccine series. Frequency distributions of time between first and second dose were generated using both open and closed claims using HealthVerity data. Separate frequency distributions were produced for patients vaccinated with mRNA-1273 and BNT162b2 and for age categories (<18, 18-64, ≥65 years). Additionally, the frequency distribution was described with all available data and before September 22, 2021, the date upon which Pfizer was granted an EUA for the original BNT162b2 formulation to be used as a booster dose for patients aged ≥65 years. A minimum of 14 days was required to occur before a second dose was captured to ensure any data noise was accounted for in this exploratory analysis.

The median time from first dose to second dose was at least 238 days for all age groups and was 22 days among pediatric patients who received a second dose prior to September 22, 2021, and 28 days among adult patients who received a second dose prior to September 22, 2021. The graphs indicate that pediatric patients had a spike at 21 days following a first dose whereas adult patients had a spike at 21 and 28 days following a first dose; the larger spike was at 28 days, regardless of the initial vaccine received.

The results suggest that many patients who have been captured as having received a heterologous primary vaccine series may be patients whose additional vaccine (second, third, fourth doses) are being captured as a primary series. For patients who do have a second vaccine event within an expected interval, a clear trend exists for patients receiving a second dose at 21 or 28 days.

The results of this exploratory analysis supported the decision to use a data-driven approach to identify a second dose. The calendar time distribution of observed vaccine doses suggests a second heterologous dose of mRNA-1273 or BNT162b2 should be defined as a dose occurring within 16 to 33 days of the first dose, aligning with the minimum and maximum intervals for a homologous dose.

**Supplementary Figure 1a: Distribution of days between dose 1 and dose 2 among patients who received a BNT162b2 first dose followed by an mRNA-1273 second dose**

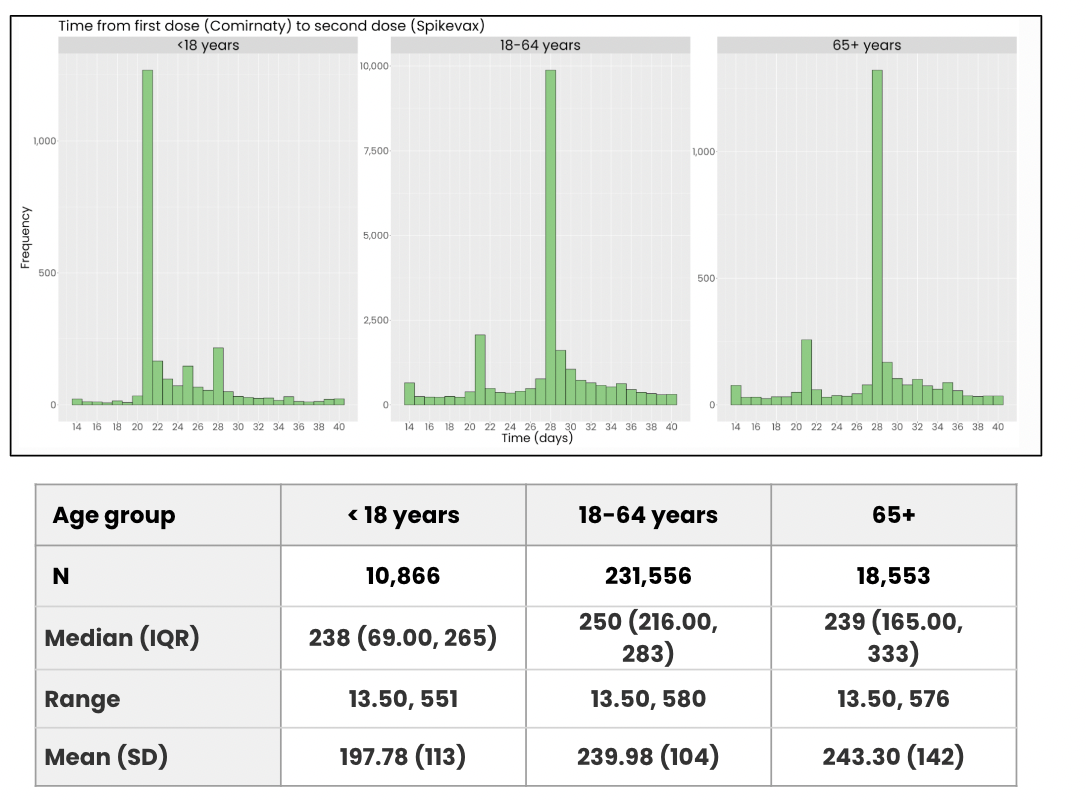

**Supplementary Figure 1b: Distribution of days between dose 1 and dose 2 among patients who received an mRNA-1273 first dose followed by a BNT162b2 second dose**

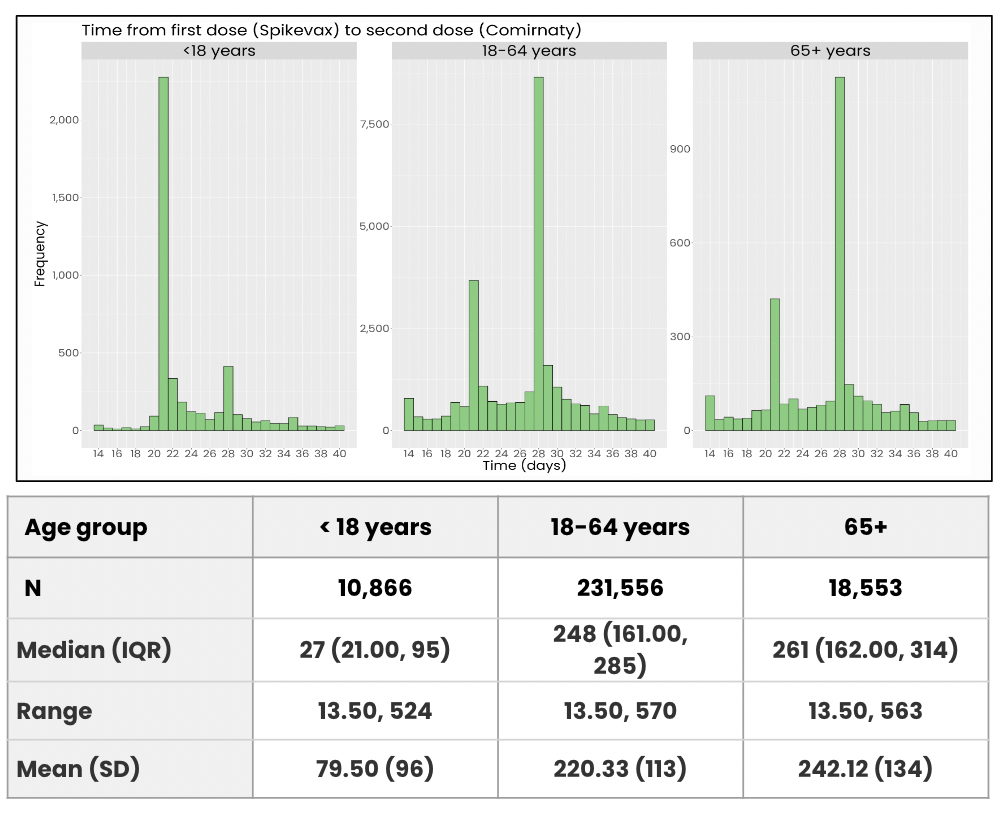

**Supplementary Figure 1c: Distribution of days between dose 1 and dose 2 among patients who received a BNT162b2 first dose followed by an mRNA-1273 second dose, where a second dose was required to occur before September 22, 2021**

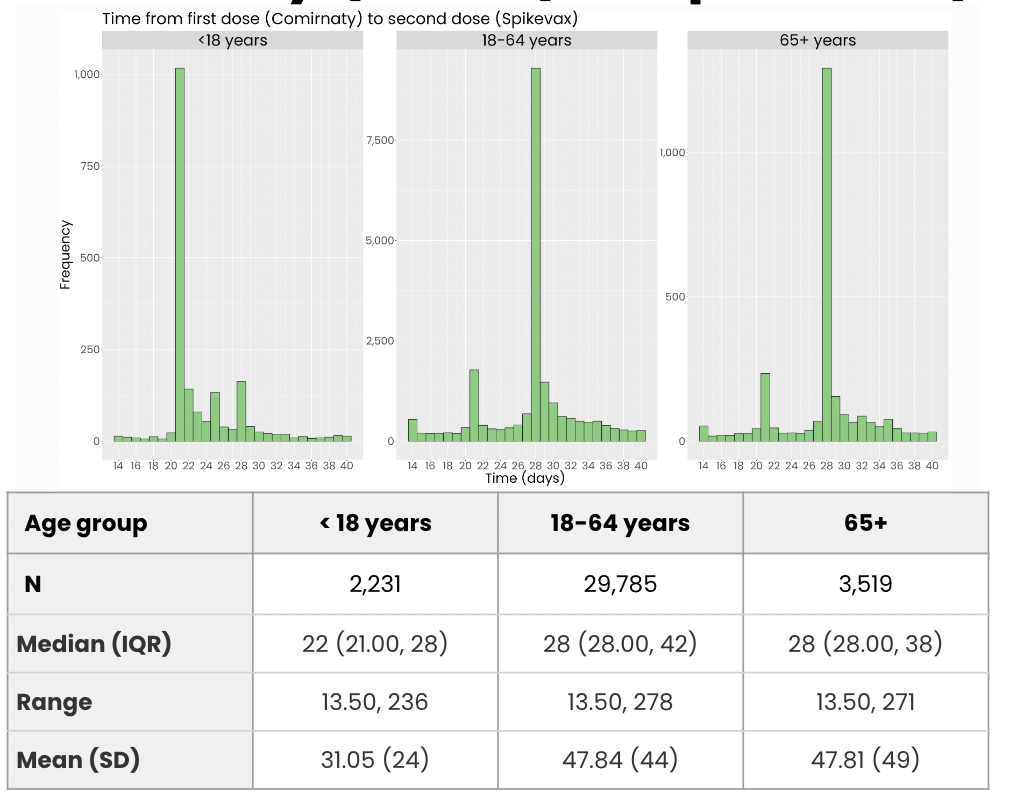

**Supplementary Figure 1d: Distribution of days between dose 1 and dose 2 among patients who received an mRNA-1273 first dose followed by a BNT162b2 second dose, where a second dose was required to occur before September 22, 2021**

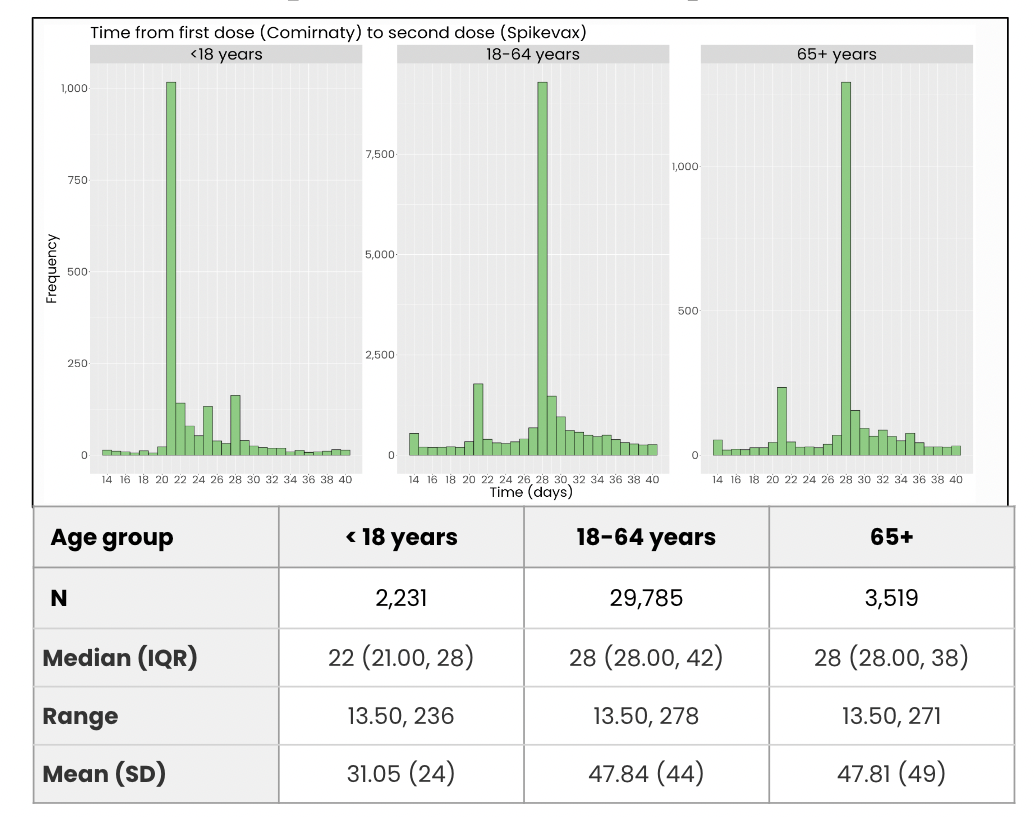

IQR, interquartile range; SD, standard deviation.

**Minimum interval between dose 2 and dose 3**

An exploratory analysis was completed to assess the minimal interval that should be applied to define a third dose following a defined second dose. Frequency distributions of time between the defined second dose and the observed third dose were generated using both open and closed claims in HealthVerity data. Separate frequency distributions were produced for patients vaccinated with mRNA-1273 and BNT162b2 and for age categories (<18, 18-64, 65+ years) and by IC status. Additionally, a booster dose was required to occur on or after September 22, 2021, based on results from the dose 1 and dose 2 exploration.

The median time between a defined second dose and an observed booster dose ranged from 210 days to 250 days across vaccine type, age groups, and IC groups. A visual inspection of the frequency distributions suggests that there were no substantial differences between IC and non-IC patients.

In the small subset of patients with <80 days between a defined second dose and an observed booster dose, there were minor peaks at 28 days among patients who received mRNA-1273 vaccines and 21 and 28 days among patients who received BNT162b2 vaccines. These spikes may represent misclassified vaccine doses. These may actually be second doses or are not booster doses.

Based on these exploratory results and incorporating both Centers for Disease Control and Prevention booster dose recommendations and the date of the BNT162b2 EUA for boosting among the elderly and IC, a third dose had to be identified on or after September 22, 2021, and had to have occurred with a minimum window of 42 days after a defined second dose, in order to increase the probability of patients indexing on a third dose that was received as a booster dose.

**Supplementary file 2. Details on defining COVID-19 hospitalization**

Unlike other administrative claims databases, there is no notion of an inpatient claim in the HealthVerity data. Inpatient claims are identified via two inpatient indicators: 1) “Inpatient indicator” is a HealthVerity-derived attribute that is true if the variable “Date Admitted” is not null in the raw data and it is ≤61 days before or after the associated “Date of Service.” It should be noted that “Date Admitted” is sparsely populated and is only present for one contributing data vendor (PS34); 2) “Suspected inpatient indicator,” an Aetion-derived indicator, is defined based on Claim Type, Place of Service, Bill Type, and Revenue Codes. A potential implication of the non-linked inpatient claims in HealthVerity is that an inpatient event may be misclassified as an outpatient event.

Another feature of the HealthVerity data is that the individual HealthVerity and Aetion-derived inpatient indicators are not combined into a single hospitalization event with start and stop dates or admission and discharge dates. Therefore, a patient could have many consecutive hospital claims on their record that correspond to a single hospital visit.

**Supplementary Figure 2: Capture of hospitalization in HealthVerity versus the real world**

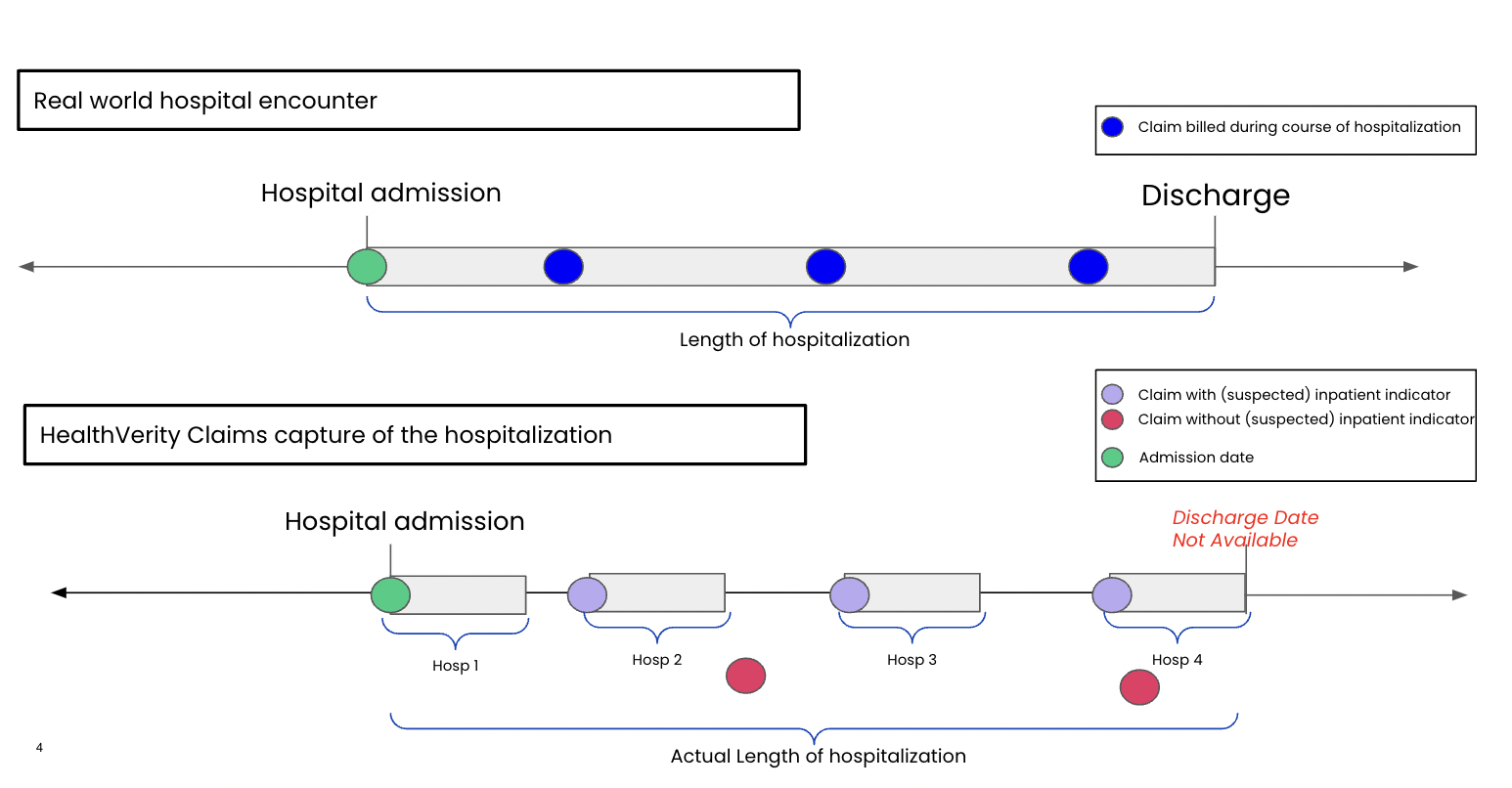

In order to capture individual diagnoses occurring during a hospitalization and to define a single hospitalization event, we considered concurrent and adjacent claims with evidence of an inpatient place of service as part of a single hospitalization episode.

Some individual inpatient claims captured in HealthVerity data occurred on non-adjacent days. For this reason, a maximum allowable gap between individual claims is applied to define a hospitalization episode. For this study, a maximum allowable gap of 4 days between adjacent inpatient claims was used. This gap allowed for the incorporation of claims that may not be reported during weekends or holidays, while allowing the capture of continuous hospitalization claims through extended holiday weekends (Example: July 4 was a Monday) and for periods in which delays in processing result in lags for which claims appear on a patient record. Restricting the allowable gap to a maximum of 4 days supports the minimization of the risk of misclassifying hospital readmissions. Furthermore, if a patient is discharged and readmitted to a hospital within a period of 4 days, we made the assumption that the primary reason for readmission was related to the primary reason for the original admission.

**Supplementary file 3. Defining respiratory distress**

To increase the specificity of the COVID-19 hospitalization algorithm, patients were required to have a COVID-19 diagnosis code and evidence of respiratory distress during a hospitalization episode. This mitigated the risk of capturing a hospitalization where a COVID-19 diagnosis code was captured as a rule-out diagnosis. Respiratory distress was identified based on code lists from a validation study performed by Kluberg et al [1] and a study utilizing EHR data to develop an algorithm to identify severe COVID-19 by Garry et al [2]. Table S3a below describes the respiratory distress definitions that were assessed in the primary and sensitivity analyses.

**Table S3a. Definition of respiratory distress assessed for the primary outcome feasibility**

|  | Included diagnoses: |
| --- | --- |
| Conditions included in the primary outcome definition | Unspecified coronavirus infection  Pneumonia due to SARS-associated coronavirus  Acute respiratory distress syndrome  Any pneumonia  Sepsis  Bronchitis  Cough  Shortness of breath  Diagnosis of indicating failed intubation  Hypoxemia  Supplemental O_2_ use  Ventilation-related diagnosis codes  Respiratory infection |
| Conditions included in the sensitivity outcome definition | Unspecified coronavirus infection  Pneumonia due to SARS-associated coronavirus  Acute respiratory distress syndrome  Any pneumonia  Sepsis |

NOS, nitric oxide synthase; SARS, severe acute respiratory syndrome.
